# Neuroanatomical-Based Machine Learning Prediction of Alzheimer’s Disease Across Sex and Age

**DOI:** 10.1101/2025.05.06.25327103

**Authors:** Bhaavin K. Jogeshwar, Senbao Lu, Benjamin C. Nephew

**Affiliations:** Worcester Polytechnic Institute, 100 Institute Rd, Worcester, 01609, Massachusetts, United States

**Keywords:** Alzheimer’s Disease, Machine Learning, Random Forest, Neuroimaging, Anatomical MRI, Sex, Age Differences, Feature Importance

## Abstract

Alzheimer’s Disease (AD) is a progressive neurodegenerative disorder characterized by cognitive decline and memory loss. In 2024, in the US alone, it affected approximately 1 in 9 people aged 65 and older, equivalent to 6.9 million individuals. Early detection and accurate AD diagnosis are crucial for improving patient outcomes. Magnetic resonance imaging (MRI) has emerged as a valuable tool for examining brain structure and identifying potential AD biomarkers. This study performs predictive analyses by employing machine learning techniques to identify key brain regions associated with AD using numerical data derived from anatomical MRI scans, going beyond standard statistical methods. Using the Random Forest Algorithm, we achieved 92.87% accuracy in detecting AD from Mild Cognitive Impairment and Cognitive Normals. Subgroup analyses across nine sex- and age-based cohorts (69–76 years, 77–84 years, and unified 69–84 years) revealed the hippocampus, amygdala, and entorhinal cortex as consistent top-rank predictors. These regions showed distinct volume reductions across age and sex groups, reflecting distinct age- and sex-related neuroanatomical patterns. For instance, younger males and females (aged 69-76) exhibited volume decreases in the right hippocampus, suggesting its importance in the early stages of AD. Older males (77-84) showed substantial volume decreases in the left inferior temporal cortex. Additionally, the left middle temporal cortex showed decreased volume in females, suggesting a potential female-specific influence, while the right entorhinal cortex may have a male-specific impact. These age-specific sex differences could inform clinical research and treatment strategies, aiding in identifying neuroanatomical markers and therapeutic targets for future clinical interventions.

## 1. Introduction

Alzheimer’s Disease (AD) is a neurodegenerative condition primarily affecting the elderly, leading to a progressive decline in cognitive abilities and impacting millions globally. As AD advances, patients increasingly require assistance with daily tasks, making it a significant health concern. In the U.S., AD is currently the fifth leading cause of death among individuals aged 65 and older [1]. This highlights an urgent need for research focused on prevention, intervention, and therapeutic strategies to manage the growing societal and familial impact of AD.

### 1.1. Prevalence of AD

The global prevalence of AD has seen a marked increase, with recent statistics showing a 147.95% rise in incidence and a 160.84% increase in prevalence from 1990 to 2019 [2]. Estimates indicate a prevalence rate of 760.5 per 100,000 individuals between 2015–2020, reflecting a substantial burden [3]. Regionally, studies from South Korea report an increase in AD incidence and prevalence among individuals aged 40 and older between 2006 and 2015 [4]. In 2023, around 6.7 million Americans aged 65 and older were estimated to be living with AD, a figure projected to reach 13.8 million by 2060 in the absence of breakthroughs in treatment [1, 5].

AD not only impacts cognitive function but is also frequently associated with Neuropsychiatric or Behavioral and Psychological Symptoms of Dementia (BPSD), affecting up to 90% of dementia patients [6]. Economically, AD presents a high burden, with an average annual per-patient cost of $20,461, with costs varying by disease stage: $14,675 for mild, $19,975 for moderate, and $29,708 for severe stages [7]. These statistics highlight the need for effective policy and intervention strategies to manage the escalating global impact of AD.

### 1.2. Early Challenges of AD

The early stages of Alzheimer’s Disease (AD) present primarily with cognitive difficulties, as brain changes related to AD may begin up to 20 years before symptoms become apparent [8]. Initial symptoms include mild memory lapses, such as forgetting names, appointments, or everyday items, which can be mistaken for typical aging. As AD progresses, cognitive challenges intensify, impacting tasks like following conversations, organizing activities, and managing finances. Memory loss extends to important events, faces, or personal history, accompanied by mood changes and difficulty with problem-solving.

In advanced AD stages, cognitive impairment becomes severe, affecting recognition of close family members and basic communication skills. Individuals often need continuous care as they lose the ability to manage self-care tasks independently. This progression is linked to the accumulation of beta-amyloid plaques and tau tangles in the brain [9, 10], which disrupt cell communication and lead to widespread neuron death, underscoring the degenerative nature of AD and the critical need for research into its underlying mechanisms [11, 12].

### 1.3. ADNI Dataset

The Alzheimer’s Disease Neuroimaging Initiative (ADNI) [13] is a longitudinal multicenter study designed to develop clinical, imaging, genetic, and biochemical biomarkers for the early detection and tracking of AD. Launched in 2004 with a $67 million investment from the National Institute on Aging and private contributors, the first phase (ADNI-1) enrolled 200 cognitively normal elderly participants, 400 with mild cognitive impairment (MCI), and 200 with early AD. Through brain scans, genetic profiles, and biomarkers from blood and cerebrospinal fluid, ADNI-1 aimed to identify precise biomarkers to enhance early diagnosis and monitor disease progression.

ADNI’s focus on structural MRI and PET scans provides valuable insights into glucose metabolism and amyloid accumulation, which are critical for understanding AD development. This initiative not only supports AD prevention, intervention, and treatment but also enables global access to its findings, advancing research across disciplines. In this study, we utilized MRI scans from AD, MCI, and cognitively normal (CN) individuals from the ADNI database, accessing raw and preprocessed data from the ADNI website to maintain consistency and support the study’s objective of exploring biomarkers related to AD progression.

### 1.4. Supervised Learning in Neuroimaging

Supervised machine learning plays a pivotal role in neuroimaging by enabling the analysis of labeled datasets to make accurate predictions or classifications. In this context, algorithms are trained on data where the outcomes are known, allowing them to identify patterns and relationships within the data. This contrasts with unsupervised learning, which works with unlabeled data to uncover inherent data structures. In neuroimaging, supervised learning techniques, such as support vector machines and random forests, are particularly valuable for analyzing MRI scans and identifying biomarkers associated with AD and other neurological conditions. These methods outperform traditional statistical techniques like linear regression or ANOVA, as they can capture nonlinear relationships and complex interactions within the data [14]. Supervised learning is also instrumental in developing predictive models for early disease detection, treatment response, and patient outcome prognosis [15, 16]. Recent advancements have emphasized its potential for early AD detection and clinical progression prediction, highlighting the integration of multimodal imaging data, such as PET, fMRI, and EEG, to create more comprehensive datasets. However, challenges remain in integrating these modalities for tracking disease progression [17]. Machine learning’s capacity to connect neuroimaging data with biological mechanisms and provide fine-scale insights into brain structure continues to advance early detection and intervention strategies for neurological diseases.

### 1.5. Subgroup Analyses in AD

Subgroup analyses are crucial in understanding the heterogeneity of AD and identifying neuroanatomical markers associated with the disease’s progression. In this study, we stratify the dataset into sex-specific and age-specific groups to examine variations in brain structure across different demographic segments. These analyses aim to uncover distinct patterns that could predict the presence of AD and enhance personalized diagnostic and therapeutic approaches. By focusing on specific subgroups, we can better understand how AD manifests differently among various populations, allowing for more targeted interventions.

#### 1.5.1. Rationale for Sex and Age-Specific Predictions

Sex differences in AD are well-documented, with females exhibiting a higher prevalence compared to males, potentially due to hormonal and genetic factors [18, 19]. At age 45, the estimated lifetime risk of developing AD is 1 in 5 for women and 1 in 10 for men [20]. In 2024, women accounted for a disproportionate number of AD cases, with 4.2 million out of 6.9 million individuals aged 65 and older diagnosed with the disease [20, 21]. These disparities highlight the importance of investigating sex-specific predictors, which may uncover underlying biological and genetic factors influencing disease progression. Similarly, age is a major risk factor for AD, with the incidence rising as individuals age, particularly after 65 [22]. Analyzing age-specific predictors can provide insights into the variations in neuroanatomical markers across different age groups, helping to identify age-related vulnerabilities and develop tailored interventions. Subgroup analyses, therefore, offer a more nuanced understanding of AD, facilitating more effective and individualized treatment strategies.

### 1.6. Future of AD

The future of AD lies in advancing early detection, preventive strategies, and personalized interventions. Research is focusing on how lifestyle factors, such as diet, physical activity, and cognitive engagement, may reduce the risk of AD onset. Early intervention, facilitated by the identification of novel biomarkers, is seen as crucial in slowing or preventing disease progression [22]. Additionally, innovative diagnostic tools and machine learning techniques hold the potential to uncover subtle neuroanatomical changes that predict AD development. As research progresses, these advancements will improve individual outcomes and alleviate the broader societal burden of AD by enabling timely interventions and more effective treatments. The collective effort to understand and address AD will empower individuals to make informed decisions regarding prevention and care, ensuring a future with reduced impact from this neurodegenerative disease.

In addition to our study, other research has explored diverse ML methodologies and datasets to advance AD diagnosis and understanding. In [23], researchers devised an ML workflow to interpret black-box models using model-agnostic Shapley values. Unlike traditional feature importance techniques, this approach provided individual explanations for each subject and examined the intricate relationships between features and predictions. The effectiveness of the workflow was assessed by training XGBoost and RF models to analyze various stages of AD. The ADNI and the Australian Imaging, Biomarker, and Lifestyle flagship study of Ageing (AIBL) cohorts were used for model training, totaling 1700 scans. Another study proposed a novel method for simultaneous differential diagnosis of CN, MCI, and AD by combining volumetric measurements, cortical thickness measurements, hippocampal texture, and hippocampal shape from structural MRI scans [24]. This approach uses linear discriminant analysis (LDA) classification to integrate multiple MRI biomarkers. The method was trained on 504 ADNI subjects and AIBL data, achieving comparable classification accuracy and area under the receiver operating characteristic curve to the Computer-Aided Diagnosis of Dementia (CADDementia) challenge. In [25], the deep learning-based FastSurfer pipeline was compared to the FreeSurfer pipeline for extracting volumetric features from MRI scans. FastSurfer demonstrated remarkably shorter execution times than FreeSurfer. The authors trained RF models on data from the ADNI cohort. They validated them on an independent test set within ADNI and a subset of the AIBL dataset, comprising 1565 scans from ADNI and 545 scans from AIBL. Results showed similar performance between models trained on FastSurfer and FreeSurfer data. The study in [26] introduces an image-fusion technique for integrating PET and T1-weighted MRI scans, enhancing feature fusion for AD detection. Additionally, ensemble classification methods, including Gradient Boosting (GB) and Support Vector Machine with Radial Basis Function (SVM RBF) for Multi-Class classification and SVM RBF + AdaBoost + GB + RF for Binary-Class classification, were proposed. The analysis included 600 scans from ADNI.

The study in [27] developed an automated ML method for classifying different stages of cognitive impairment, using cortical thickness measurements from 1167 MRI scans and achieving an overall accuracy of 75%. The authors in [28] presented a multi-modality classification framework using RF classifiers, integrating MRI volumes, FDG-PET signals, CSF biomarkers, and genetic data to achieve classification accuracies of 89% for AD vs. HC and 75% for MCI vs. HC. In [29], the authors presented the efficacy of RF classifiers using structural MRI measures, achieving a sensitivity/specificity of 88.6%/92.0% in distinguishing AD from HC, with improved sensitivity/specificity for predicting MCI-to-AD conversion when including demographic and genetic data. The study in [30] focused on predicting MCI-to-AD conversion using an RF model trained on clinical data from 383 early MCI patients, resulting in an accuracy of 93.6%. Finally, the authors in [31] tested RF models on 2250 MRI scans, achieving high performance (90.2%), and identified the hippocampus, amygdala, and inferior lateral ventricle as major contributors to classification accuracy.

Our study builds upon these methodologies by integrating the Random Forest Algorithm with various cross-validation techniques to predict AD and detect neuroanatomical markers of AD. We achieved an accuracy of 92.87% and an F1 score of 92.84% on our dataset, demonstrating robust predictive performance. These diverse approaches highlight the ongoing advancements in ML methodologies and their application to neuroimaging data, demonstrating the potential to substantially enhance AD diagnosis and understanding.

In this report, we use the ADNI dataset to extract brain MRI scans, which are then parcellated into various brain regions for analysis. The focus is on identifying brain regions that are most relevant in predicting AD across different demographic subgroups. We have covered the following topics. Section 2 shows the methodology. Section 3 presents the results and analysis of the study. Section 4 discusses and concludes the paper with our overall findings.

## 2. Methodology

### 2.1. ADNI Dataset

Our study utilized a dataset of 815 structural MRI scans, comprising 281 Cognitive Normal (CN), 332 Mild Cognitive Impairment (MCI), and 202 Alzheimer’s disease (AD) scans, from 344 subjects aged 69 to 84 years. We applied preprocessing techniques including Multiplanar Reconstruction (MPR), GradWarp, B1 Correction, N3, and Scaling. The scans were T1-weighted Magnetization-Prepared Rapid Gradient-Echo (MPRAGE) images from the first phase of the Alzheimer’s Disease Neuroimaging Initiative (ADNI-1).

Subjects also completed the Mini-Mental State Exam (MMSE) [32] and Clinical Dementia Rating (CDR) [33] tests, typically administered around the time of imaging. The MMSE assesses cognitive function across several domains, including orientation to time and place, registration, attention and calculation, recall, language, repetition, and the ability to follow complex commands and write a sentence. The CDR, based on informant reports, stages the overall severity of dementia. These tools are critical in Alzheimer’s research, providing standardized, quantifiable measures of cognitive decline over time. MMSE and CDR scores were considered when selecting subjects for this study. Table 1 outlines the demographic characteristics of the ADNI dataset used.

**Table 1:** ADNI demographic characteristics.

|  | **N** | **Sex (F:M)** | **Age (years)** | **MMSE** | **CDR** |
| --- | --- | --- | --- | --- | --- |
| **CN** | 281 | 171:110 | 73.7-81.0 | 29-30 | 0 |
| **MCI** | 332 | 104:228 | 71.6-83.0 | 25.4-29 | 0.5 |
| **AD** | 202 | 108:94 | 69.4-83.6 | 16.0-25.0 | 0.5-1.0 |
|  | **815** | **383:432** | **69.4-83.6** |  |  |

We divided the dataset into subgroups based on sex and age, covering male and female subjects across two age ranges: 69–76 years and 77–84 years. The subgroups are as follows:

1. Both male and female subjects aged 69 to 84
2. Male-only subjects aged 69 to 84
3. Female-only subjects aged 69 to 84
4. Both male and female subjects aged 69 to 76
5. Male-only subjects aged 69 to 76
6. Female-only subjects aged 69 to 76
7. Both male and female subjects aged 77 to 84
8. Male-only subjects aged 77 to 84
9. Female-only subjects aged 77 to 84

Table 2 presents the distribution of diagnoses and sample sizes within each sub-group.

**Table 2:** Distribution of ADNI Participants by Age, Sex, and Diagnosis.

| **Age Group**<br>(in years) | **N** |  |  |  |
| --- | --- | --- | --- | --- |
|  | **Group** | **Both sexes**<br>(M+F) | **Males**<br>(M) | **Females**<br>(F) |
| Ages 69-84 | CN | 281 | 110 | 171 |
|  | MCI | 332 | 228 | 104 |
|  | AD | 202 | 94 | 108 |
|  |  | **815** | **432** | **383** |
| Ages 69-76 | CN | 117 | 51 | 66 |
|  | MCI | 151 | 112 | 39 |
|  | AD | 106 | 39 | 67 |
|  |  | **374** | **202** | **172** |
| Ages 77-84 | CN | 164 | 59 | 105 |
|  | MCI | 181 | 116 | 65 |
|  | AD | 96 | 55 | 41 |
|  |  | **441** | **230** | **211** |

### 2.2. FastSurfer

For this study, we used FastSurfer, a deep learning-based neuroimaging pipeline, for volumetric analysis of brain MRI scans [34]. FastSurfer significantly reduces processing time compared to traditional methods like FreeSurfer, while maintaining accuracy.

We used FastSurfer’s `--seg_only` command to analyze brain volumes from 815 MRI scans. On our lab machine (Ubuntu 22.04, 64GB RAM, NVIDIA GeForce RTX 2080 SUPER), each scan took approximately 1 minute for volumetric analysis, compared to around 3 hours per scan with FreeSurfer. This speed made FastSurfer the optimal choice for handling large datasets in this study.

Our analysis targeted volume changes in both cortical and subcortical regions to assess structural differences associated with age, sex, and AD. Focusing on these volume measurements ensured consistency across brain regions, allowing us to identify relevant patterns for the study.

During preprocessing, we encountered two challenges. First, an internal parcellation error in FastSurfer affected cerebellar segmentation. As the cerebellum was not central to our study, we excluded it from analysis using the `--no_cereb` flag. Second, one MRI scan exhibited poor contrast and missing brain regions, which led to an IndexError during processing. This scan was removed from the dataset to maintain data quality. A visual comparison between the discarded and retained scan is shown in Figure 1.

**Figure 1.**
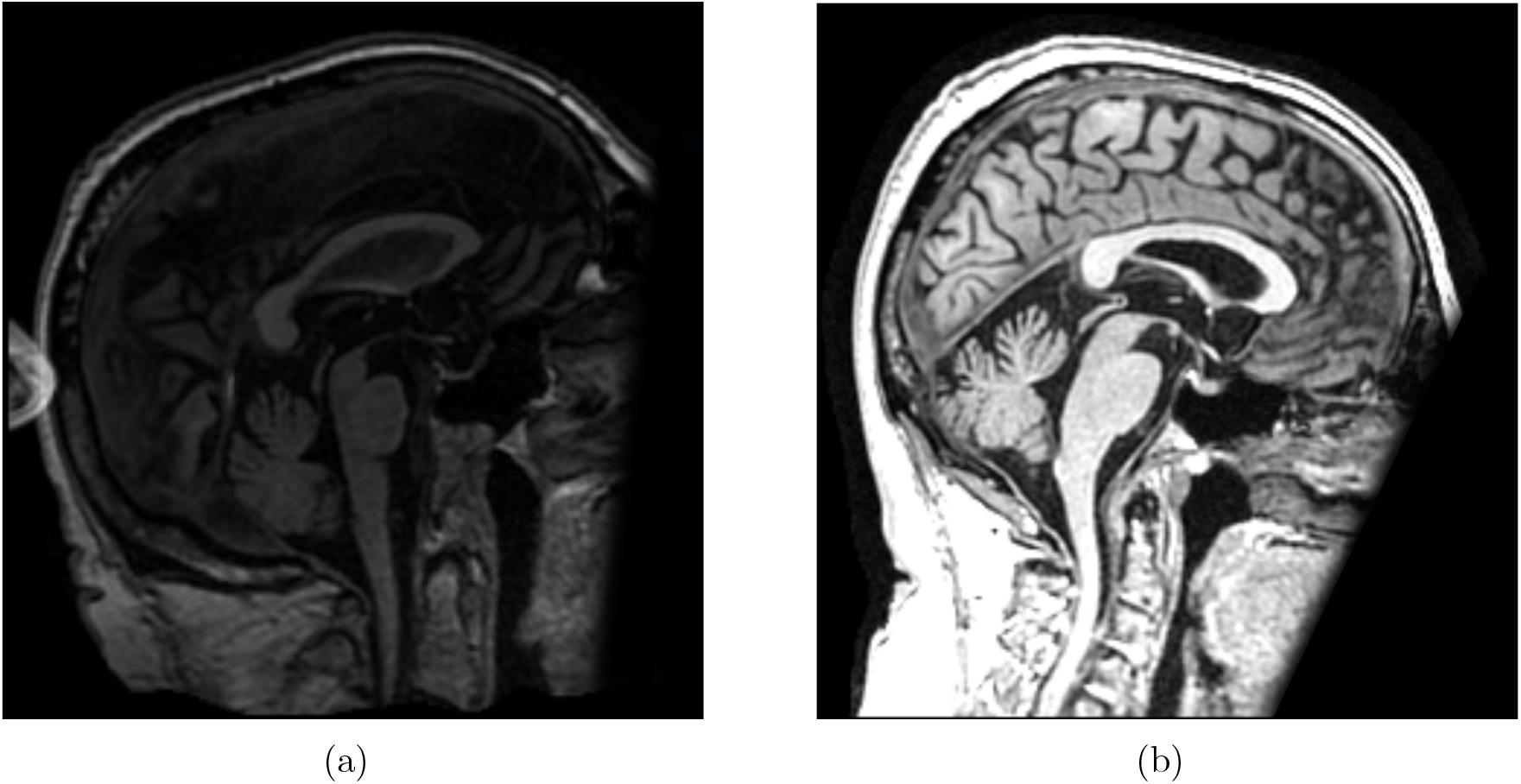
Comparison of a discarded scan with a high-quality scan. (a) Defaced scan with missing brain regions that triggered a processing error. (b) High-quality scan processed successfully.

### 2.3. Class Imbalance

Class imbalance, where one class is significantly underrepresented, can lead to biased models favoring the majority class. To address this, we employed the Synthetic Minority Over-sampling Technique (SMOTE) from the `imblearn` library [35]. SMOTE generates synthetic samples by interpolating between minority class instances, helping balance the dataset for improved performance in predicting minority classes. Although other techniques like ADASYN [36] are available, we chose SMOTE for its demonstrated superior performance in comparable scenarios [37].

After applying normalization and splitting the data into training and testing sets, SMOTE was used to balance the training set for random forest classifiers. This improved the model’s performance, particularly in handling class imbalance, with accuracy increasing from 86-88% to 90-93%.

### 2.4. Classification Model: Random Forest

Random Forest (RF) is an ensemble learning algorithm commonly used for classification and regression tasks [38]. It builds multiple decision trees on bootstrapped samples of the dataset, considering random subsets of features at each split. The final prediction is made by aggregating the outputs of all trees, either through majority voting (classification) or averaging (regression). RF also ranks features by assessing their contribution to reducing impurity or error across the trees. In this study, we implemented RF for Alzheimer’s Disease prediction, as outlined in Figure 2. We adapted the machine learning pipeline from [39], originally developed for suicide risk prediction, to classify AD using neuroanatomical MRI data. The primary objective is to identify the brain regions with strong predictive power for distinguishing Alzheimer’s Disease from other diagnostic groups based on MRI-derived features.

**Figure 2.**
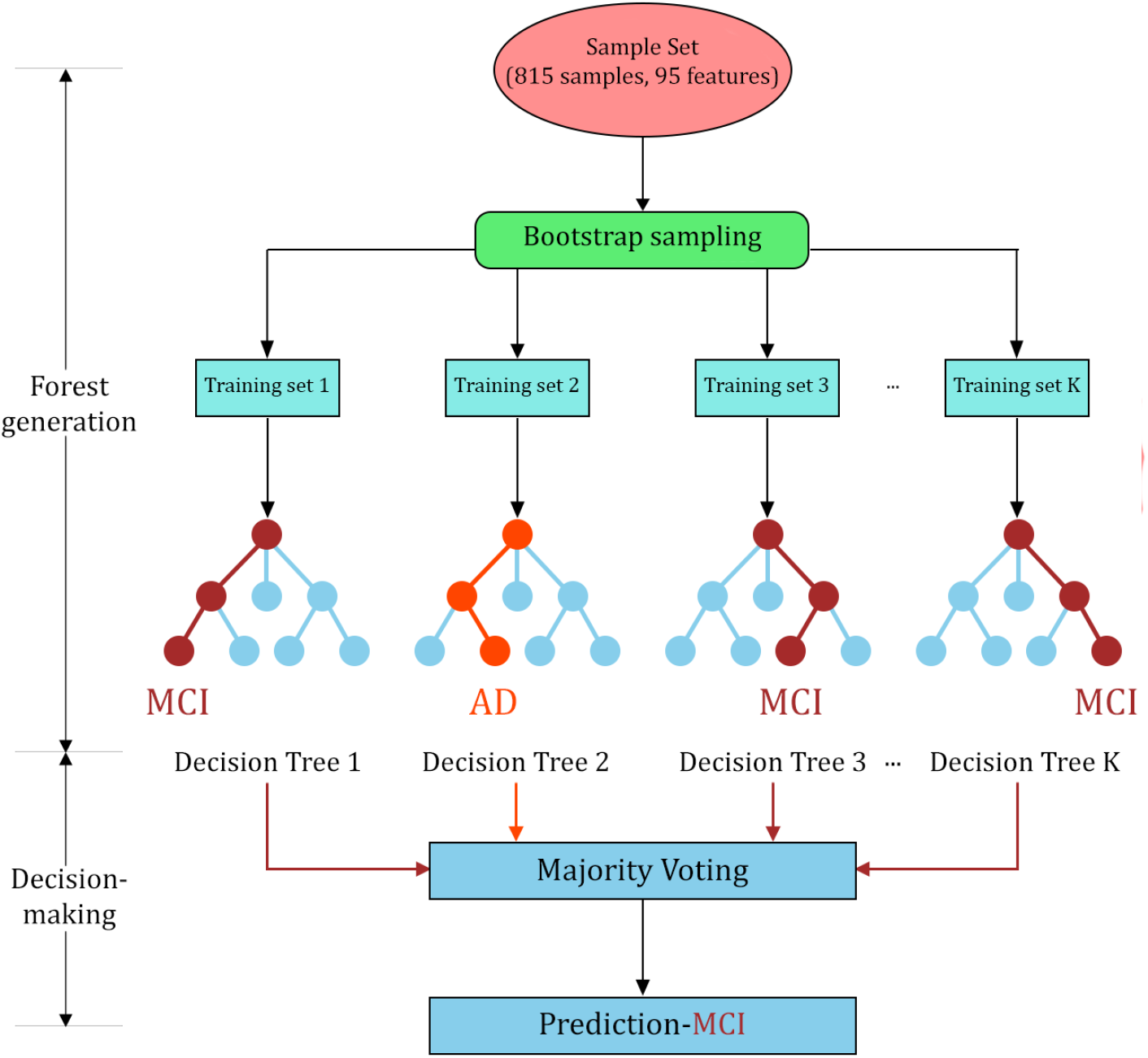
An example flowchart of Random Forest classifier.

Hyperparameter tuning was performed using cross-validation. The number of estimators varied between *{*100, 200, 300*}*, and maximum tree depths of *{*None, 10, 20, 30*}* were tested to balance model complexity and generalization. Additionally, ‘min_samples_split’ and ‘min_samples_leaf’ parameters were tuned to regularize the model. The final configuration was selected based on accuracy and area under the ROC curve (AUC), ensuring robust generalization performance across demographic subgroups.

### 2.5. RF-Based Neuroanatomical Marker Analysis

A key advantage of the Random Forest algorithm is its ability to identify the top contributing features and assess their importance in classification. In this study, we used the model’s built-in feature importance metric to rank brain regions based on their predictive relevance for AD. This ranking enabled us to pinpoint which neuroanatomical structures most strongly influenced the model’s predictions.

### 2.6. Model Validation

This section outlines the validation techniques employed to assess the performance of the RF models for predicting AD.

#### 2.6.1. K-Fold Cross-Validation

K-fold cross-validation divides the dataset into *k* equal parts (folds), where each fold serves as the validation set once, and the remaining *k −* 1 folds are used for training. This process is repeated *k* times to ensure each fold is used for validation, and the performance is averaged across all iterations. For this study, we used 5-fold cross-validation (*k* = 5), providing a balanced approach for estimating model performance metrics, such as accuracy, precision, recall, and F1-score. This method helps to minimize bias and variance in the performance estimates, leading to more reliable model evaluation.

#### 2.6.2. Stratified K-Fold Cross-Validation

Stratified K-Fold Cross-Validation preserves the class distribution in each fold, ensuring that each fold is representative of the overall dataset’s class distribution. This method prevents underrepresentation of smaller classes within any fold. In this study, we employed 5-fold Stratified Cross-Validation (*k* = 5), ensuring balanced class representation across all folds for more reliable model evaluation.

#### 2.6.3. Leave-One-Out Cross-Validation

In Leave-One-Out Cross-Validation (LOOCV), each sample in the dataset is used once as a validation set, while the remaining samples form the training set [39]. This process is repeated for all samples, resulting in *n* folds, where *n* is the number of samples. LOOCV offers a thorough assessment of model performance by maximizing the use of available data. We applied LOOCV in this study to further evaluate model stability and robustness.

#### 2.6.4. Implementation

In our implementation, we use the `cross_val_predict` function from the `sklearn.model_selection` module to perform K-Fold, Stratified K-Fold, and Leave-one-out cross-validation. This function efficiently handles the splitting of the dataset and evaluation of the model across multiple folds.

### 2.7. Performance Metrics

To evaluate the effectiveness of the RF classifier in predicting AD, we used the follwoing performance metrics: accuracy, precision, recall, and F1-score. These metrics provide a comprehensive assessment of model performance across different aspects of classification.

1. Accuracy
  - Accuracy measures the proportion of correctly classified instances out of the total. It is particularly reliable when the dataset has balanced class distributions but may be less informative for imbalanced datasets.
2. Precision
  - Precision measures the model’s ability to correctly identify true positives from all predicted positives. It is calculated as the ratio of true positives to the sum of true and false positives. It is especially useful when minimizing false positives is critical.
3. Recall
  - Also known as sensitivity, recall quantifies the model’s ability to correctly identify all actual positives. It is the ratio of true positives to the sum of true positives and false negatives, prioritizing minimizing false negatives.
4. F1-score
  - The F1-Score is the harmonic mean of precision and recall, balancing both metrics and proving useful when both false positives and false negatives are significant.

These metrics were computed using functions from the `sklearn.metrics` module, such as `accuracy_score`, `precision_score`, `recall_score`, and `f1_score`.

In summary, we used the ADNI dataset for MRI scans and conducted volumetric analysis using FastSurfer. The dataset was first divided by sex to explore gender-specific differences in AD progression and then further split into two age groups to investigate age-related variations. An RF classifier, combined with SMOTE, and three validation techniques, was employed to identify key brain regions associated with AD. Performance metrics were evaluated, and the following chapter presents the results.

## 3. Results

### 3.1. FastSurfer outputs

The preprocessed MRI scans from ADNI were parcellated into 100 brain regions using FastSurfer’s `--seg-only` pipeline, based on the Desikan-Killiany-Tourville (DKT) Atlas. These regions included 31 cortical areas per hemisphere, 33 subcortical regions, and five areas of the corpus callosum. Regions with zero volumes were excluded, leaving 95 regions for analysis. A list of these 95 regions is provided in the Supplementary section (Chapter 5. Figure 3 shows a CN subject’s brain parcellation in Freeview neuroimaging software.

**Figure 3.**
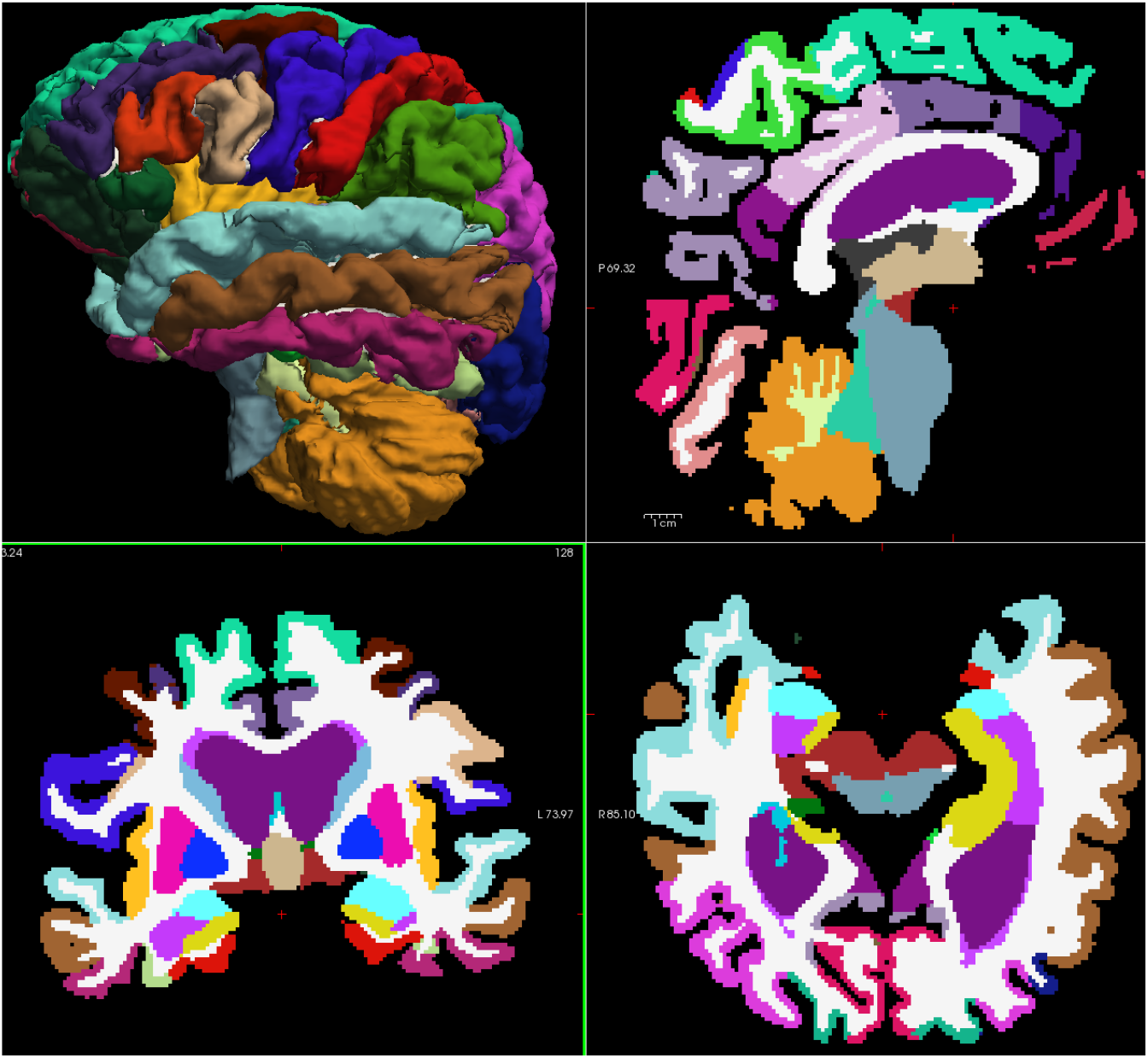
Brain parcellation in Freeview.

### 3.2. Feature Engineering

We normalized the data to improve model performance. Specifically, we introduced a custom normalization approach: each brain region’s volume was normalized by the total volume of all 95 brain regions, ensuring consistent feature scaling between 0 and 1, as shown in Equation (1):

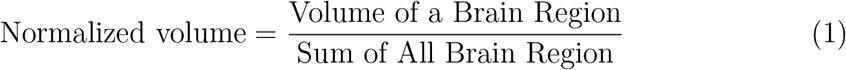

This custom normalization led to a slight improvement in data quality, reflected in our model’s performance. Using RF with this custom normalization strategy, we achieved an accuracy of **0.883**, compared to **0.877** without normalization. We also explored traditional methods, such as `MinMaxScaler()` from `sklearn.preprocessing`, which produced comparable accuracy, supporting the effectiveness of both approaches. Given these results, we prioritized custom normalization in further analyses.

After data normalization, we organized the dataset into subgroups based on sex and age. These subgroups included male, female, and combined subjects, further divided into two age ranges: 69-76 and 77-84. To address class imbalance, we applied the `SMOTE` technique. For hyperparameter optimization, we explored Randomized Search and BayesSearchCV, both yielding similar results, but ultimately chose the ParameterGrid approach for its slight performance advantage. We fine-tuned Random Forest parameters using hyperparameter tuning and validated them through K-fold, stratified K-fold, and leave-one-out cross-validation. The most influential features were then identified across all subgroups.

### 3.3. Subgroup Analyses

#### 3.3.1. Statistical Analysis of Brain Regions

Several brain regions were analyzed for statistical comparisons. Figure 4 shows the average volumes of the left and right hippocampus, amygdala, and entorhinal cortex in CN, MCI, and AD patients. The statistical differences among the three groups were assessed using a two-sample Z-test for each pair of brain regions. Significant differences (p *<* 0.01) were found across CN vs. MCI, MCI vs. AD, and CN vs. AD comparisons. The Z-test was chosen to demonstrate significant differences in brain region volumes, which were influenced by the consistently low p-values, emphasizing the reliability of our statistical outcomes.

**Figure 4.**
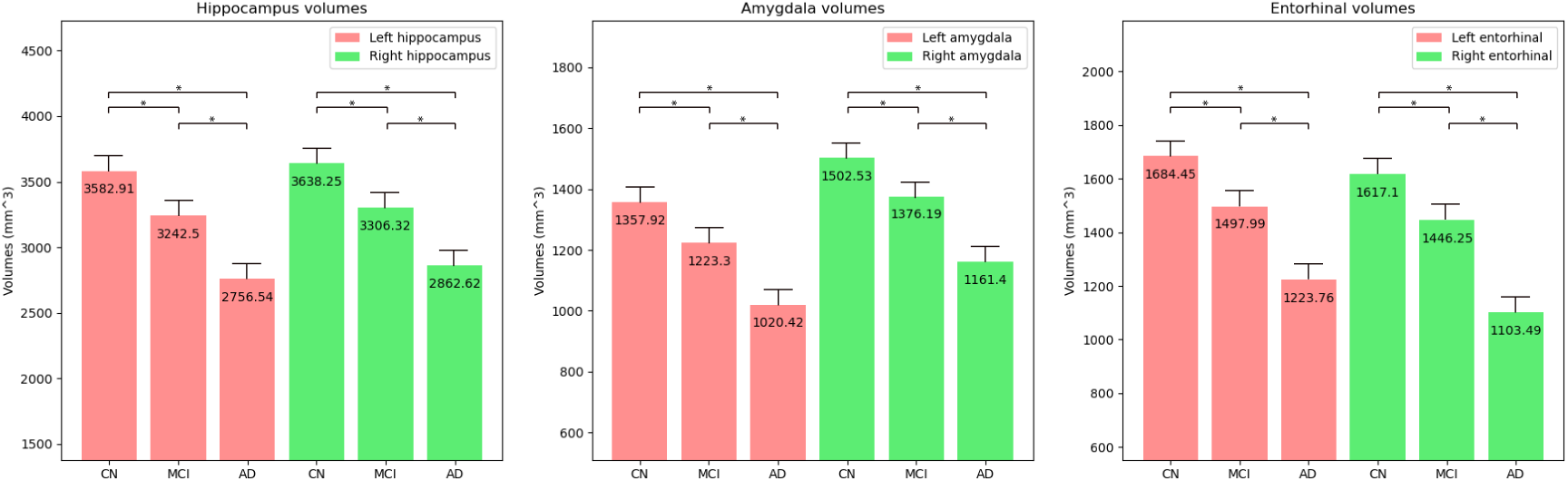
Average volumes of left and right hippocampus, amygdala, and entorhinal cortex in CN, MCI, and AD subjects, illustrating atrophy across cognitive states. Statistical significance is marked with * (p *<* 0.01).

Figure 5 presents the average volumes of four critical brain regions, Left-Inf-Lat-Vent (LILV), left inferior parietal cortex, left inferior temporal cortex, and left middle temporal cortex, across cognitive states in both sexes aged 69-84. Significant differences (p *<* 0.01) were observed in all comparisons except for CN to MCI in the left inferior parietal and left inferior temporal cortices. These results highlight the impact of widespread brain atrophy in AD, with certain areas showing early volume changes and others exhibiting more pronounced differences as the disease progresses.

**Figure 5.**
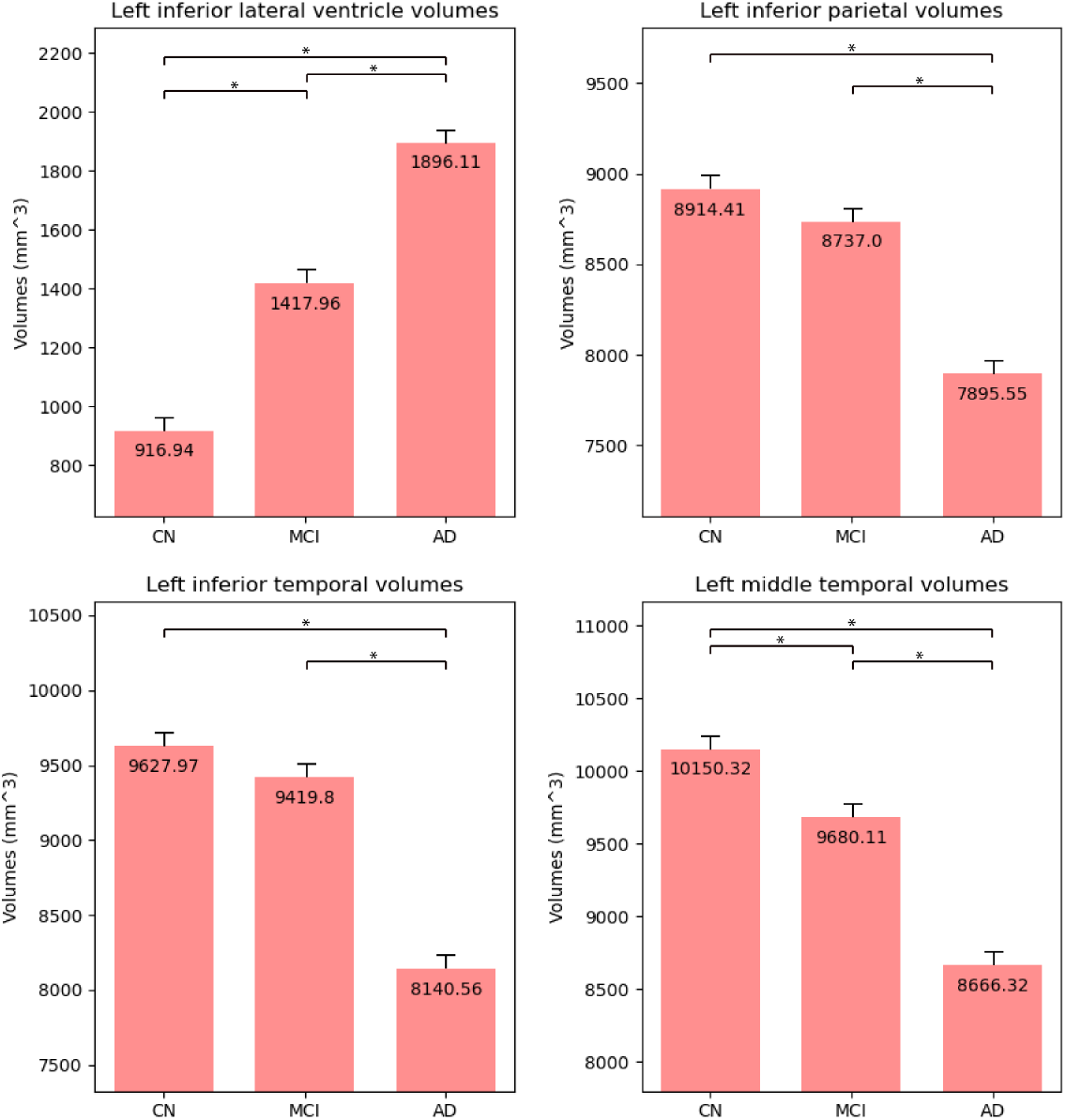
Average volumes of Left Inferior Lateral Ventricle, left inferior parietal cortex, left inferior temporal cortex, and left middle temporal cortex in CN, MCI, and AD subjects, illustrating the trend of ventricular enlargement and cortical atrophy across cognitive states. Statistical significance is marked with * (p *<* 0.01).

#### 3.3.2. Understanding the Benjamini-Hochberg Correction

When comparing brain region volumes across groups (CN, MCI, and AD), the risk of false positives increases due to multiple statistical tests. The Benjamini-Hochberg (BH) correction addresses this by adjusting the p-values to control the false discovery rate [40].

Initially, p-values from the tests indicate the likelihood of results occurring by chance, but they don’t account for multiple comparisons. After applying the BH correction, the adjusted p-values provide a more accurate assessment of the true significance in brain region volume differences among the groups.

### Comparison of P-Values

#### After Benjamini-Hochberg Correction

Table 3 presents the p-values before applying the Benjamini-Hochberg correction, while Table 4 shows the p-values after adjustment. These tables enable a comparison of significance levels before and after correcting for multiple comparisons. The BH-corrected p-values offer a more reliable indication of true significance in brain region volume differences among the groups. While most comparisons remained highly significant, regions such as the left inferior parietal cortex and left inferior temporal cortex showed non-significant differences between CN and MCI groups after correction. This suggests that early atrophy in these regions may be less distinct during the mild cognitive impairment stage, emphasizing the importance of focusing on brain regions with consistently significant differences across diagnostic groups.

**Table 3:** P-values before Benjamini-Hochberg correction.

Comparison of P-Values
| Brain Region | CN vs MCI | CN vs AD | MCI vs AD | MCI vs CN | AD vs CN | AD vs MCI |
| --- | --- | --- | --- | --- | --- | --- |
| Left-Hippocampus | 5.43e-22 | 1.24e-92 | 5.43e-22 | 1.05e-30 | 1.24e-92 | 1.05e-30 |
| Right-Hippocampus | 6.82e-17 | 1.05e-68 | 6.82e-17 | 1.91e-23 | 1.05e-68 | 1.91e-23 |
| Left-Amygdala | 1.85e-13 | 6.97e-70 | 1.85e-13 | 9.47e-26 | 6.97e-70 | 9.47e-26 |
| Right-Amygdala | 6.16e-09 | 2.65e-47 | 6.16e-09 | 1.66e-19 | 2.65e-47 | 1.66e-19 |
| ctx-lh-entorhinal | 3.92e-12 | 3.76e-56 | 3.92e-12 | 1.41e-18 | 3.76e-56 | 1.41e-18 |
| ctx-rh-entorhinal | 3.44e-09 | 2.77e-69 | 3.44e-09 | 3.39e-29 | 2.77e-69 | 3.39e-29 |
| Left-Inf-Lat-Vent | 5.09e-20 | 1.91e-27 | 6.84e-07 | 5.09e-20 | 1.91e-27 | 6.84e-07 |
| ctx-lh-inferiorparietal | 0.136 | 1.73e-15 | 1.60e-09 | 0.136 | 1.73e-15 | 1.60e-09 |
| ctx-lh-inferiortemporal | 0.0793 | 7.32e-30 | 1.74e-20 | 0.0793 | 7.32e-30 | 1.74e-20 |
| ctx-lh-middletemporal | 2.96e-04 | 8.68e-24 | 8.28e-12 | 2.96e-04 | 8.68e-24 | 8.28e-12 |

**Table 4:** P-values after Benjamini-Hochberg correction.

| Brain Region | CN vs MCI | CN vs AD | MCI vs AD | MCI vs CN | AD vs CN | AD vs MCI |
| --- | --- | --- | --- | --- | --- | --- |
| Left-Hippocampus | 8.89e-22 | 2.24e-91 | 8.89e-22 | 2.69e-30 | 2.24e-91 | 2.69e-30 |
| Right-Hippocampus | 8.77e-17 | 4.71e-68 | 8.77e-17 | 3.44e-23 | 4.71e-68 | 3.44e-23 |
| Left-Amygdala | 2.22e-13 | 6.27e-69 | 2.22e-13 | 1.89e-25 | 6.27e-69 | 1.89e-25 |
| Right-Amygdala | 6.16e-09 | 7.95e-47 | 6.16e-09 | 2.50e-19 | 7.95e-47 | 2.50e-19 |
| ctx-lh-entorhinal | 4.41e-12 | 1.36e-55 | 4.41e-12 | 1.95e-18 | 1.36e-55 | 1.95e-18 |
| ctx-rh-entorhinal | 3.64e-09 | 1.66e-68 | 3.64e-09 | 7.63e-29 | 1.66e-68 | 7.63e-29 |
| Left-Inf-Lat-Vent | 9.54e-20 | 5.72e-27 | 9.54e-20 | 7.60e-07 | 5.72e-27 | 7.60e-07 |
| ctx-lh-inferiorparietal | 0.136 | 2.60e-15 | 0.136 | 1.99e-09 | 2.60e-15 | 1.99e-09 |
| ctx-lh-inferiortemporal | 0.0821 | 2.75e-29 | 0.0821 | 3.48e-20 | 2.75e-29 | 3.48e-20 |
| ctx-lh-middletemporal | 3.17e-04 | 2.17e-23 | 3.17e-04 | 1.08e-11 | 2.17e-23 | 1.08e-11 |

#### 3.3.3. Performance Metrics

Tables 5, 6, and 7 display the aggregated performance metrics achieved through K-fold, stratified K-fold, and leave-one-out cross-validation techniques across the nine subgroups. All three methods show comparable accuracy, consistently in the low 90s. However, leave-one-out cross-validation demonstrates slightly better performance. The total execution time differs significantly, with leave-one-out taking the longest at 23.73 minutes, while the other two methods finish in just 1 minute each.

**Table 5:**
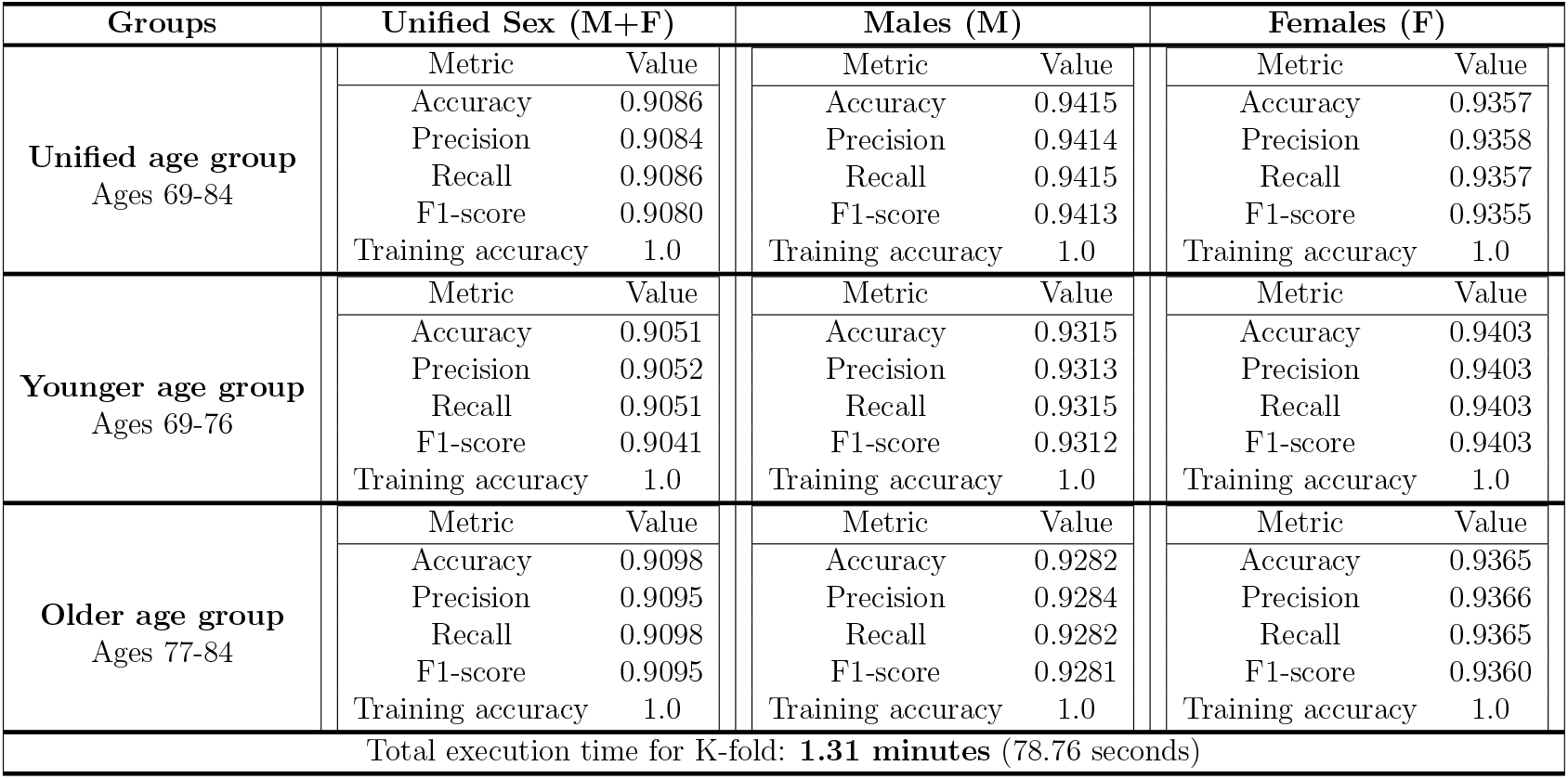
Comparison of Aggregated Performance Metrics using K-fold Cross-Validation.

**Table 6:** Comparison of Aggregated Performance Metrics using Stratified K-fold Cross-Validation.

| Groups | Unified Sex (M+F) |  | Males (M) |  | Females (F) |  |
| --- | --- | --- | --- | --- | --- | --- |
| **Unified age group**<br>Ages 69-84 | Metric | Value | Metric | Value | Metric | Value |
|  | Accuracy | 0.9187 | Accuracy | 0.9503 | Accuracy | 0.9357 |
|  | Precision | 0.9190 | Precision | 0.9502 | Precision | 0.9359 |
|  | Recall | 0.9187 | Recall | 0.9503 | Recall | 0.9357 |
|  | F1-score | 0.9178 | F1-score | 0.9502 | F1-score | 0.9355 |
|  | Training accuracy | 1.0 | Training accuracy | 1.0 | Training accuracy | 1.0 |
| **Younger age group**<br>Ages 69-76 | Metric | Value | Metric | Value | Metric | Value |
|  | Accuracy | 0.8962 | Accuracy | 0.9583 | Accuracy | 0.9353 |
|  | Precision | 0.8962 | Precision | 0.9583 | Precision | 0.9355 |
|  | Recall | 0.8962 | Recall | 0.9583 | Recall | 0.9353 |
|  | F1-score | 0.8957 | F1-score | 0.9582 | F1-score | 0.9351 |
|  | Training accuracy | 1.0 | Training accuracy | 1.0 | Training accuracy | 1.0 |
| **Older age group**<br>Ages 77-84 | Metric | Value | Metric | Value | Metric | Value |
|  | Accuracy | 0.9116 | Accuracy | 0.9224 | Accuracy | 0.9365 |
|  | Precision | 0.9123 | Precision | 0.9222 | Precision | 0.9363 |
|  | Recall | 0.9116 | Recall | 0.9224 | Recall | 0.9365 |
|  | F1-score | 0.9107 | F1-score | 0.9222 | F1-score | 0.9363 |
|  | Training accuracy | 1.0 | Training accuracy | 1.0 | Training accuracy | 1.0 |
| Total execution time for Stratified K-fold: **1.34 minutes** (80.67 seconds) |  |  |  |  |  |  |

**Table 7:** Comparison of Aggregated Performance Metrics using Leave-One-Out Cross-Validation.

| Groups | Unified Sex (M+F) |  | Males (M) |  | Females (F) |  |
| --- | --- | --- | --- | --- | --- | --- |
| **Unified age group**<br>Ages 69-84 | Metric | Value | Metric | Value | Metric | Value |
|  | Accuracy | 0.9287 | Accuracy | 0.9547 | Accuracy | 0.9435 |
|  | Precision | 0.9284 | Precision | 0.9547 | Precision | 0.9435 |
|  | Recall | 0.9287 | Recall | 0.9547 | Recall | 0.9435 |
|  | F1-score | 0.9284 | F1-score | 0.9546 | F1-score | 0.9434 |
|  | Training accuracy | 1.0 | Training accuracy | 1.0 | Training accuracy | 1.0 |
| **Younger age group**<br>Ages 69-76 | Metric | Value | Metric | Value | Metric | Value |
|  | Accuracy | 0.9205 | Accuracy | 0.9435 | Accuracy | 0.9303 |
|  | Precision | 0.9212 | Precision | 0.9435 | Precision | 0.9307 |
|  | Recall | 0.9205 | Recall | 0.9435 | Recall | 0.9303 |
|  | F1-score | 0.9200 | F1-score | 0.9430 | F1-score | 0.9304 |
|  | Training accuracy | 1.0 | Training accuracy | 1.0 | Training accuracy | 1.0 |
| **Older age group**<br>Ages 77-84 | Metric | Value | Metric | Value | Metric | Value |
|  | Accuracy | 0.9263 | Accuracy | 0.9397 | Accuracy | 0.9460 |
|  | Precision | 0.9272 | Precision | 0.9396 | Precision | 0.9477 |
|  | Recall | 0.9263 | Recall | 0.9397 | Recall | 0.9460 |
|  | F1-score | 0.9257 | F1-score | 0.9396 | F1-score | 0.9455 |
|  | Training accuracy | 1.0 | Training accuracy | 1.0 | Training accuracy | 1.0 |
| Total execution time for Leave-One-Out: **23.73 minutes** |  |  |  |  |  |  |

#### 3.3.4. RF-Based Neuroanatomical Marker Analysis

The tables 8, 9, and 10 detail the top six brain regions most frequently ranked as high-importance features across nine subgroups, identified through K-fold, stratified K-fold, and leave-one-out cross-validation techniques. These tables reveal consistent brain regions recurring as top contributors across the subgroups.

**Table 8:**
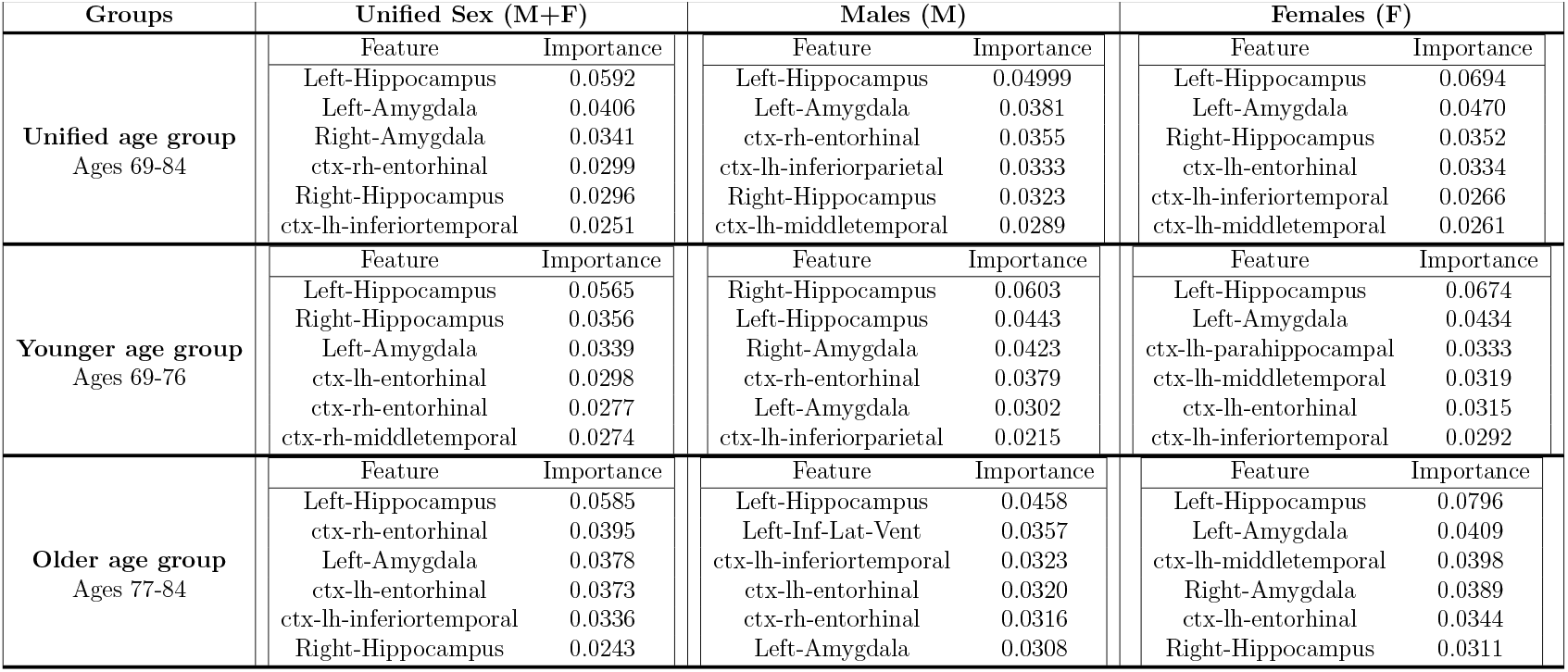
Comparison of Top Contributing Features using K-fold Cross-Validation.

**Table 9:**
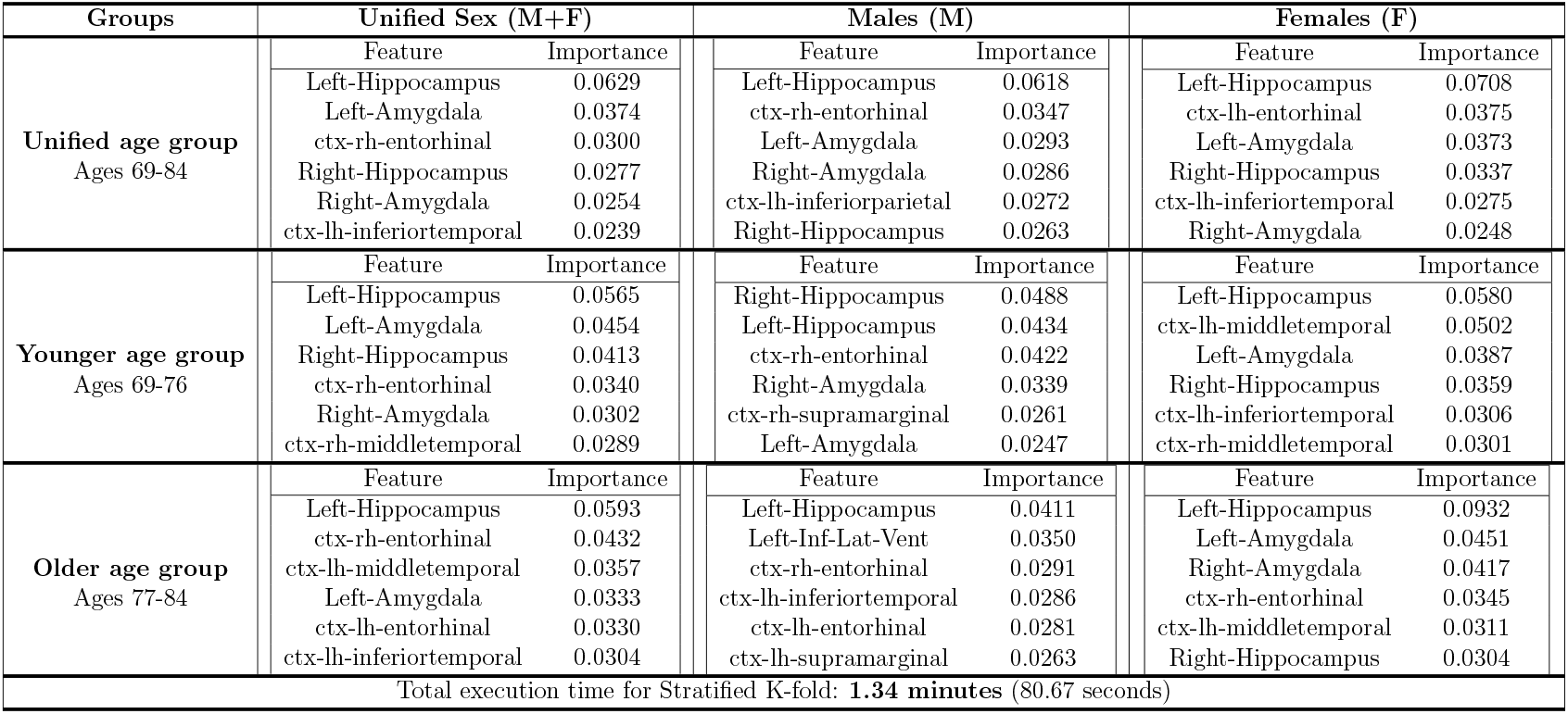
Comparison of Top Contributing Features using Stratified K-fold Cross-Validation.

**Table 10:**
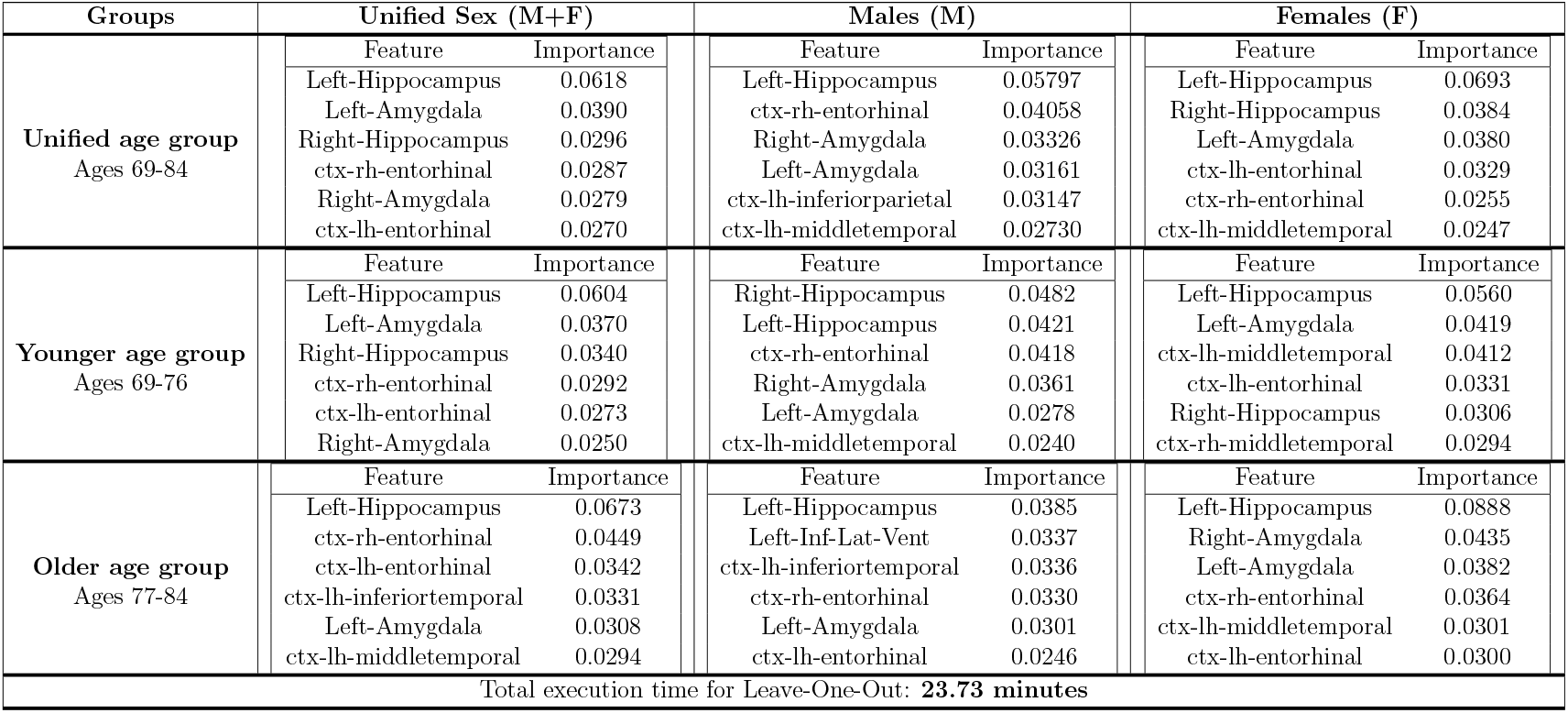
Comparison of Top Contributing Features using Leave-One-Out Cross-Validation.

To assess the consistency of feature identification, we examined the overlap of top contributing brain regions across all three validation methods. Among the 54 features analyzed, derived from the top six features in each of the nine subgroups, 39 were consistently identified across all tables, yielding a consistency score of 72.22%.

This high consistency underscores the dataset’s influence on determining the top contributing features rather than the specific validation techniques, enhancing confidence in the predictive models. Table 11 presents the 39 consistently identified features across all subgroups and validation techniques. Detailed performance metrics and top contributing features for each subgroup are provided in the Supplementary section (Chapter (5)).

**Table 11:** Consistent Top Contributing Features using K-fold, Stratified K-fold, and Leave-One-Out Cross-Validation.

| Groups | Unified Sex (M+F) | Males (M) | Females (F) |
| --- | --- | --- | --- |
| **Unified age group**<br>Ages 69-84 | Left-Hippocampus<br>Left-Amygdala<br>Right-Amygdala<br>ctx-rh-entorhinal<br>Right-Hippocampus | Left-Hippocampus<br>Left-Amygdala<br>ctx-rh-entorhinal<br>ctx-lh-inferiorparietal | Left-Hippocampus<br>Left-Amygdala<br>Right-Hippocampus<br>ctx-lh-entorhinal |
| **Younger age group**<br>Ages 69-76 | Left-Hippocampus<br>Right-Hippocampus<br>Left-Amygdala<br>ctx-rh-entorhinal | Right-Hippocampus<br>Left-Hippocampus<br>Right-Amygdala<br>ctx-rh-entorhinal<br>Left-Amygdala | Left-Hippocampus<br>Left-Amygdala<br>ctx-lh-middletemporal |
| **Older age group**<br>Ages 77-84 | Left-Hippocampus<br>ctx-rh-entorhinal<br>Left-Amygdala<br>ctx-lh-entorhinal<br>ctx-lh-inferiortemporal | Left-Hippocampus<br>Left-Inf-Lat-Vent<br>ctx-lh-inferiortemporal<br>ctx-lh-entorhinal<br>ctx-rh-entorhinal | Left-Hippocampus<br>Left-Amygdala<br>ctx-lh-middletemporal<br>Right-Amygdala |

**Table 12:**
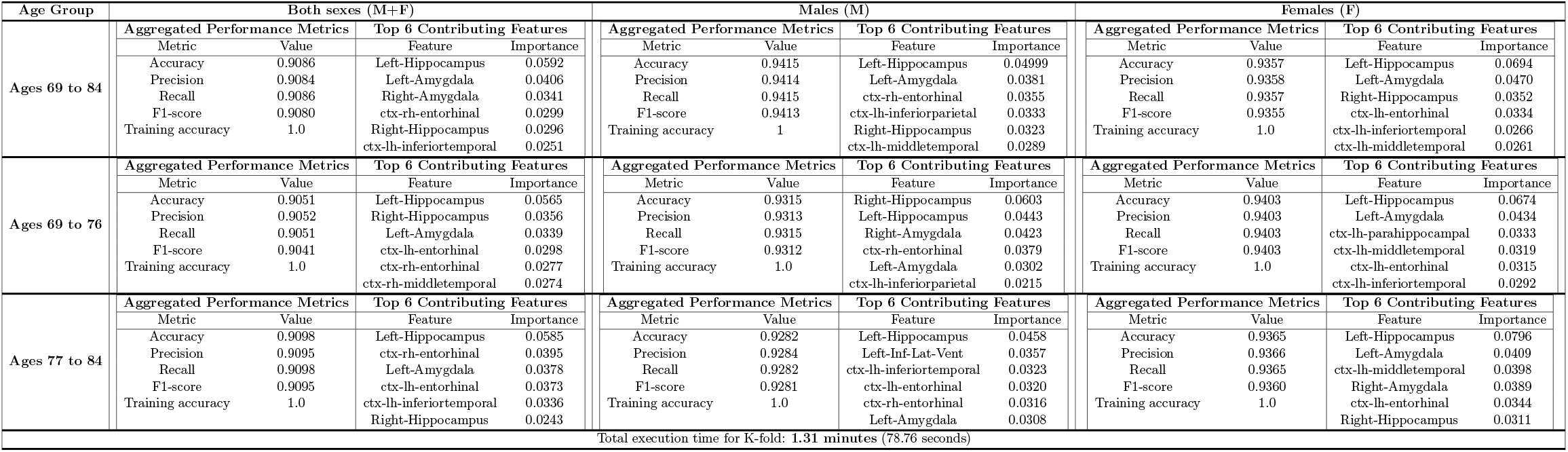
Comparison of Performance Metrics and Top Contributing Features using K-fold Cross-Validation.

An analysis of Table 11 reveals consistent patterns of brain region involvement across age and sex groups. For instance, the left hippocampus consistently decreases in volume in both males and females across all age groups. Additionally, younger males (aged 69-76) and older females (aged 77-84) exhibit substantial volume reductions in the right amygdala. The younger (69-76) age group shows substantial volume decreases in the right hippocampus, highlighting its importance in the early stages of AD across the sexes. Furthermore, older males (aged 77-84) exhibit notable volume reductions in the left inferior temporal cortex region. These findings emphasize variations in brain region predictors based on age and sex.

#### 3.3.5. Neuroanatomical Trends in AD

In studying Table 11, the neuroanatomical trends in AD indicate consistent volume reductions across all brain regions, except for the lateral ventricle, which enlarges. These trends highlight sex-specific, age-related, and regional influences on AD.

### Sex-Specific Trends

- **Similarities observed in both males and females:**
  - The left hippocampus and left amygdala are key predictors in the Unified age group (69-84).
- **Differences observed across**
  - Unified age group (69-84):
    - The ctx-lh-entorhinal (left entorhinal cortex) is a prominent predictor in females, while the ctx-lh-inferiorparietal (left inferior parietal cortex) is a prominent predictor in males.
  - Younger age group (69-76):
    - The ctx-rh-entorhinal (right entorhinal cortex) is a prominent predictor in males.
  - Older age group (77-84):
    - The ctx-lh-inferiortemporal (left inferior temporal cortex) is a leading predictor in males, while the ctx-lh-middletemporal (left middle temporal cortex) is a leading predictor in females.

### Age-Related Trends

- **Similarities Observed in both younger (69-76) and older (77-84) age groups**
  - The left hippocampus, left amygdala, and right entorhinal cortex are prominent predictors in the Unified Sex group (M+F).
  - The right entorhinal cortex is a prominent predictor in males, while the left middle temporal cortex shows prominence in females.
- **Differences Observed across**
  - Unified Sex group (M+F):
    - The left inferior temporal cortex is a key predictor in the older age group (77-84).
  - Males only:
    - The left entorhinal cortex is a prominent predictor in the older age group (77-84).
    - The right amygdala and right hippocampus are key predictors in the younger age group (69-76).
  - Females only:
    - The right amygdala is a prominent predictor in the older age group (77-84).

### Distinctive Regional Trends

- Across both sexes and age groups (69-76 and 77-84), the left hippocampus and left amygdala appear more frequently among the top-ranked features, implying their substantial role as key contributors to AD.
- The left middle temporal cortex is a prominent predictor among females across both younger (69-76) and older (77-84) age groups, suggesting a potential female-specific influence.
- Influence of the right entorhinal cortex is observed predominantly in males across both younger (69-76) and older (77-84) age groups, suggesting a potential male-specific influence.
- The right amygdala and right hippocampus are prominent predictors in younger males (aged 69-76) but not in the older male group (aged 77-84). Conversely, the right amygdala exhibits the opposite trend in females, being relevant in the older female group (aged 77-84) but not in the younger female group (aged 69-76). This observation suggests a potential interaction between age and sex affecting the right amygdala region.
- Influence of the left inferior temporal cortex and Left-Inf-Lat-Vent is male-specific in the older age group (77-84).
- The left entorhinal cortex is a prominent predictor in females and is also observed in older males (77-84).

The left inferior lateral ventricle (LILV), a cerebrospinal fluid-filled cavity that maintains brain homeostasis and cushions brain structures, often enlarges due to brain atrophy or neurodegeneration as surrounding tissues shrink. This enlargement across cognitive states is evident in Figure 5.

Figure 5 shows a marked increase in LILV volume from CN to AD, reflecting ventricular enlargement with disease progression. In contrast, the left inferior parietal, inferior temporal, and middle temporal cortices exhibit decreasing volume trends, indicating cortical atrophy as cognitive impairment advances. Similar volume reductions are seen in the hippocampus, amygdala, and entorhinal cortex (Figure 4). The left inferior parietal and temporal cortices show moderate volume loss from CN to MCI and more pronounced reductions from MCI to AD, while the left middle temporal cortex exhibits a consistent decline across all groups. These reductions point to neurodegeneration affecting regions critical for spatial awareness, visual perception, language comprehension, and memory integration.

#### 3.3.6. Insights from Visual Representations

##### Comprehensive Overview

We define feature “ranking” based on RF feature importance scores. For each of the nine subgroups, the top six features were selected based on their relative importance. These subgroup-specific rankings are detailed in Tables 8, 9, and 10. Figure 6 visualizes how frequently each brain region appeared among these top-ranked features across the subgroup analyses and cross-validation methods. This bar plot offers a comprehensive view of the consistency and prominence of brain regions in AD classification.

**Figure 6.**
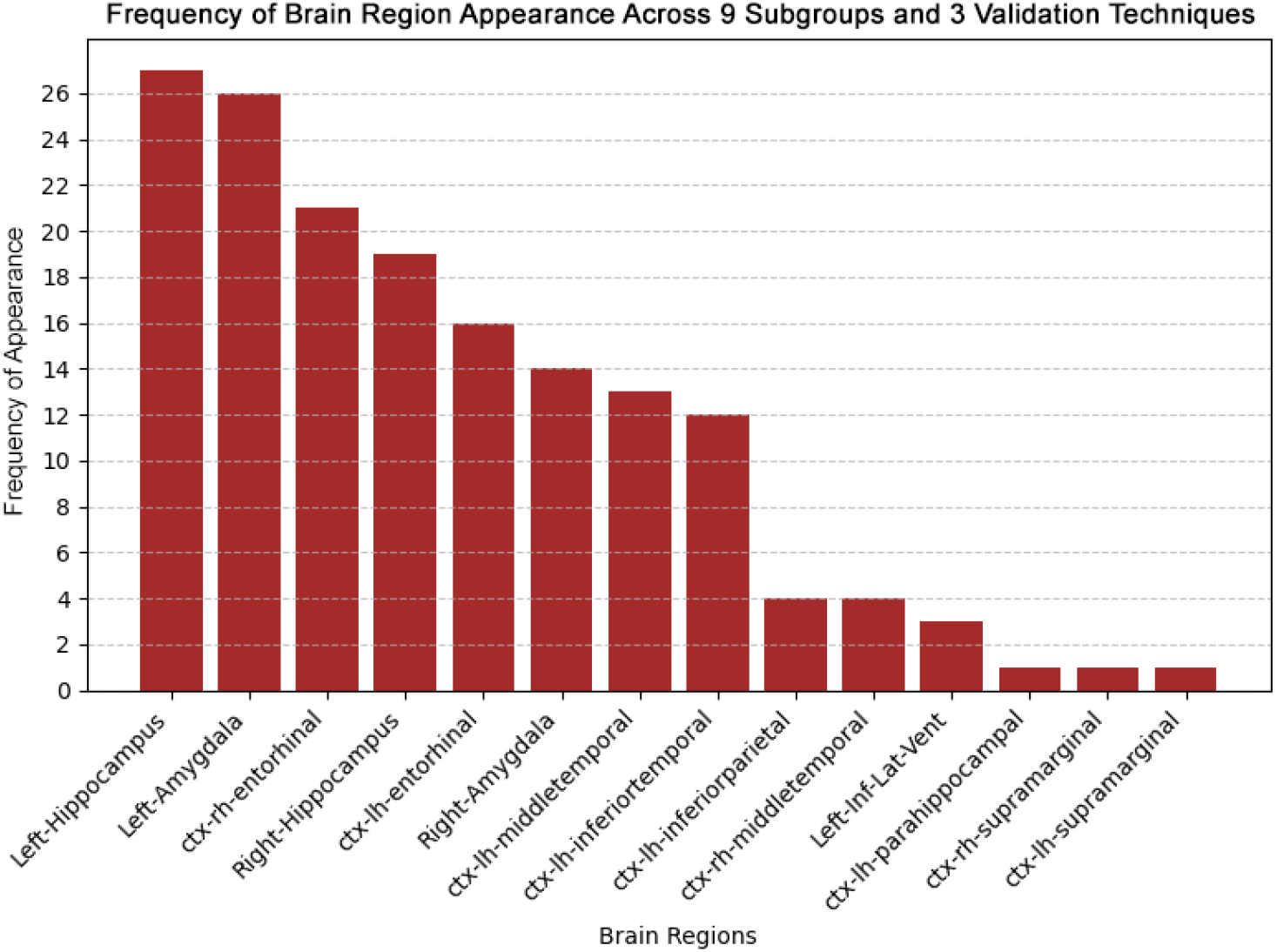
Frequency of appearance of brain regions among the top-ranked features across nine subgroups and three cross-validation techniques. Frequency of appearance denotes how often a brain region was selected among the top-ranked features across subgroup-specific models.

##### Prominence of Frequently Selected Brain Regions

Focusing on the top six brain regions with the highest frequency of appearance (Figure 7), we observe that the left hippocampus prominently features across all subgroups and validation techniques. Overall, the hippocampus, amygdala, and entorhinal cortex emerge as prominent predictive features, consistently associated with decreased volume in Alzheimer’s Disease. These regions are visualized on Freeview in Figure 8.

**Figure 7.**
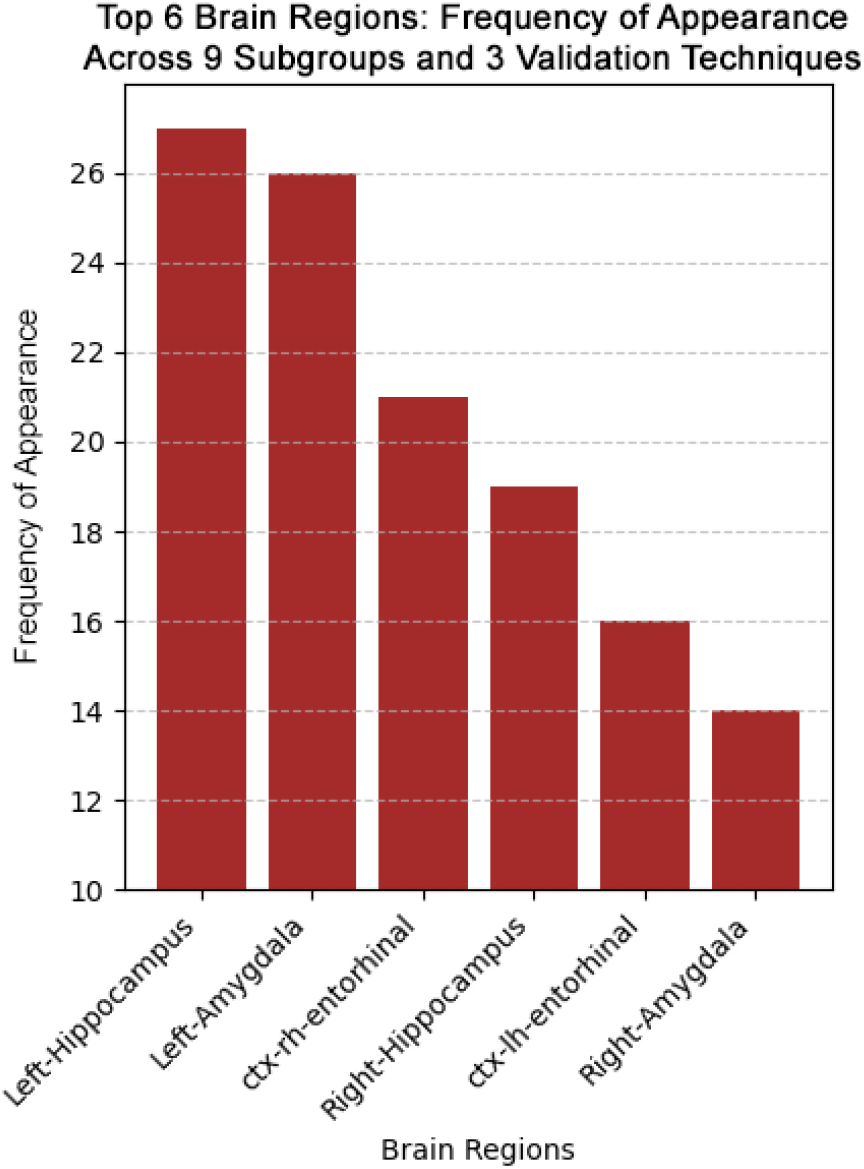
Frequency of appearance of the top six brain regions across nine subgroups and three cross-validation techniques.

**Figure 8.**
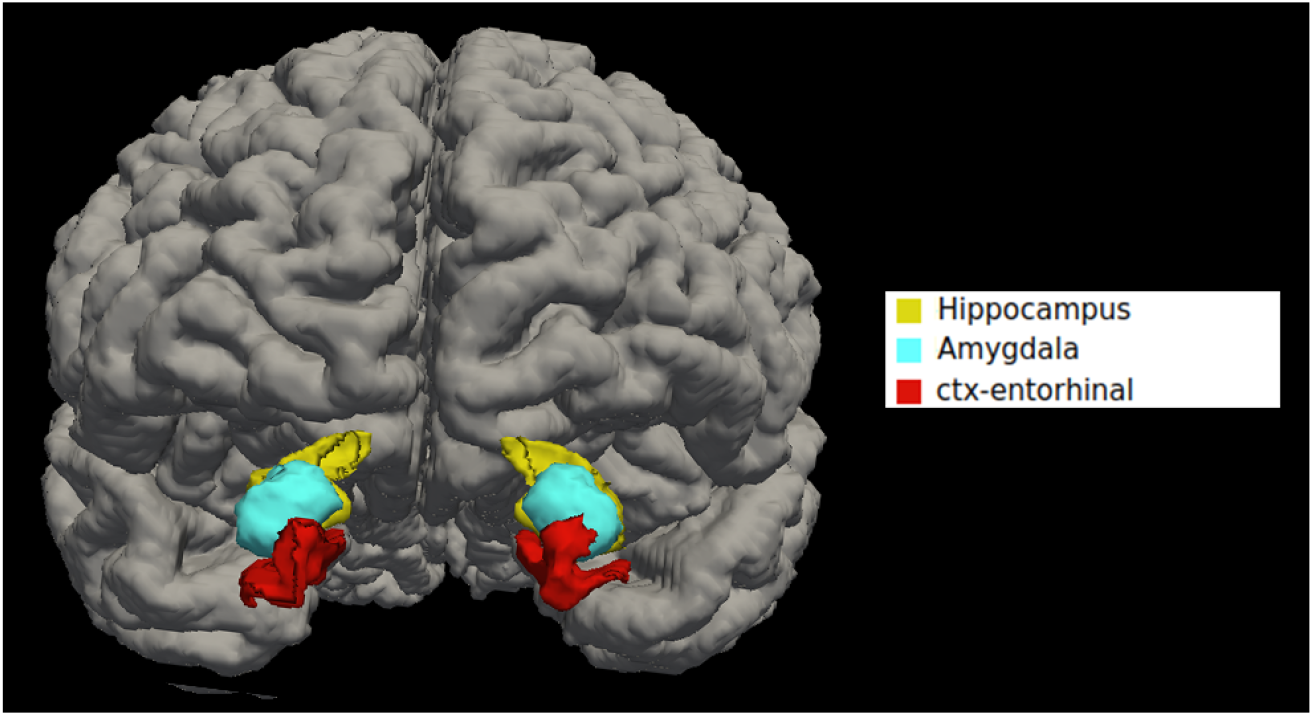
Visualization of top 6 brain regions - the hippocampus, amygdala, and entorhinal cortex on Freeview.

## 4. Discussion

The primary objective of this study was to predict CN, MCI, and AD diagnoses using neuroanatomical data from structural MRI scans in sex- and age-specific groups. Using the Random Forest (RF) algorithm with three cross-validation techniques, we achieved accuracies of 0.9086 for K-fold CV, 0.9187 for Stratified K-fold CV, and 0.9287 for LOOCV. Key brain regions identified through regression analysis, i.e., hippocampus, amygdala, and entorhinal cortex, consistently showed reduced volumes in AD across all demographic groups. Sex- and age-specific patterns of structural changes were observed, supporting the development of personalized diagnostic and therapeutic strategies. These findings highlight the potential for tailored treatment plans that consider unique neuroanatomical profiles, ultimately improving patient outcomes.

### 4.1. Potential neuroanatomical markers of MCI and AD

We aimed to identify structurally affected brain regions in individuals aged 69-84 to capture the impact of AD across this age range. By including cross-sectional scans of CN, MCI, and AD patients, rather than focusing solely on CN and AD data, we detected structural changes across disease stages. This approach highlighted brain regions where changes emerge or diminish from CN to MCI and MCI to AD, offering a more comprehensive perspective. The top six features, i.e., left and right hippocampus, amygdala, and entorhinal cortex, were identified as the most relevant markers for AD, and their roles in the disease will be further explored.

#### 4.1.1. Role of hippocampus and amygdala

Our analysis consistently identified the left hippocampus and left amygdala as key regions contributing to AD across both sexes and age groups (69-76 and 77-84). In younger males (69-76), the right hippocampus and right amygdala were also significant predictors, though their influence diminished in older males (77-84). In contrast, the right amygdala emerged as relevant in older females (77-84), but not in younger females, suggesting potential sex and age-related interactions in AD progression.

#### 4.1.2. The hippocampus

The hippocampus is essential for learning and memory, with its sub-regions contributing to the formation of episodic memory [41, 42]. In the initial stages of AD, rapid tissue loss occurs in the hippocampus, leading to functional disconnection from other brain regions [43]. This damage disrupts the hippocampus’s ability to function correctly, leading to memory loss and cognitive decline, which are hallmark symptoms of AD. Additionally, the hippocampus is involved in various cognitive functions beyond memory, such as emotional regulation and spatial orientation, and its dysfunction contributes to the broader cognitive impairment observed in AD.

#### 4.1.3. The amygdala

The amygdala, integral to the limbic system, is linked to emotional disorders like autism, anxiety disorders, and AD, particularly in the context of fear. In the early stages of AD, both the amygdala and hippocampus change, leading to personality changes and emotional abnormalities like anxiety, mania, and irritability [44]. Because of early amygdala damage, mild stages of AD often exhibit neuropsychiatric symptoms. Eventually, about 80% of AD patients experience symptoms like hallucinations, delusions, paranoia, anxiety, agitation, and mood disturbances as the disease progresses [45, 46].

#### 4.1.4. Role of entorhinal cortex

Our analysis identified distinct sex- and age-specific trends in the entorhinal cortex. The right entorhinal cortex consistently showed volume reduction in both younger (69-76) and older (77-84) males, but this pattern was not observed in younger females. This suggests a male-specific influence in the right entorhinal cortex. In contrast, the left entorhinal cortex exhibited volume reduction in females across the unified age group (69-84) and only in older males (77-84). These findings highlight potential interactions between sex and age in the entorhinal cortex’s involvement in AD progression.

The entorhinal cortex serves as an initial focal point for the deposition of plaques and tangles in AD [47]. Plaques are abnormal clusters of beta-amyloid protein fragments that accumulate between nerve cells in the brain, while tau tangles are twisted tau protein fibers that form inside the brain cells. AD is characterized by these distinctive brain abnormalities, amyloid-*β* plaques and tau protein neurofibrillary tangles, which actively influence the neurodegenerative process [48]. During the early stages of AD, there is a buildup of tau protein within the entorhinal cortex, which propagates to the hippocampus [49]. Autopsy-based anatomical and histological investigations of Alzheimer’s-afflicted brains have shown a progressive pattern of neurodegeneration, commencing in the second layer of the entorhinal cortex and gradually advancing to encompass the hippocampus, temporal cortex, frontoparietal cortex, and subcortical nuclei. The entorhinal cortex in individuals with AD experiences impaired function in processing sensory information and transmitting it for memory consolidation [50, 51]. This impairment results in substantial memory decline and difficulties with spatial navigation.

Our results provide detailed insights into the neurodegenerative patterns of AD, particularly highlighting volume reductions in the hippocampus, amygdala, and entorhinal cortex, areas critical for memory and emotional regulation. These findings align with previous studies while also revealing sex- and age-specific variations in neurodegeneration. By identifying these key predictors, our study contributes to a deeper understanding of how AD affects different demographics and advances knowledge of its pathology.

### 4.2. Sex and Age Subgroup Analyses

Sex-based differences play a significant role in the prevalence and progression of AD. In 2024, nearly two-thirds of Americans with Alzheimer’s are women; of the 6.9 million people aged 65 and older with the disease, 4.2 million are women, and 2.7 million are men [20, 21]. Additionally, the lifetime risk of developing AD at age 45 is about 20% for women and 10% for men [20]. Males tend to experience slower structural loss but faster age-related brain volume decline compared to females. This pattern corresponds with the distinct regional influences observed in AD, such as the male-specific prominence of the right entorhinal cortex and the female-specific influence of the left middle temporal region, which may help explain these sex-based differences.

Men, with their larger head size and cerebral brain volume compared to women, approximately 10% larger [52], may be less vulnerable to AD-related pathological factors, experiencing slower structural loss [53]. However, they experience faster agerelated brain volume decline compared to women [54]. Despite having similar levels of AD pathology, women are more likely to receive a clinical diagnosis than men [55]. Upon AD diagnosis, men often display reduced atrophy in various brain regions [56]. Women tend to have a higher proportion of gray matter, whereas men have a higher percentage of white matter [57]. These differences are often attributed to sex chromosomes and sex hormones, although the precise mechanism by which sex hormones affect brain structures remains unclear [58, 53].

As of 2024, the prevalence of Alzheimer’s dementia in the U.S. increases with age: 5.0% of individuals aged 65 to 74, 13.2% of those aged 75 to 84, and 33.4% of people aged 85 and older are affected [20, 21]. Annually, 4 out of 1,000 individuals aged 65 to 74, 32 out of 1,000 aged 75 to 84, and 76 out of 1,000 aged 85 and older develop AD [59]. Additionally, an estimated 8% to 11% of Americans aged 65 and older, approximately 5 to 7 million, may experience MCI due to AD [60]. The risk of developing AD sharply increases with age, which aligns with our study’s identification of age-specific neuroanatomical contributors, such as the male-specific involvement of the left entorhinal cortex in the older age group (77-84). Most AD cases are diagnosed after age 65, with prevalence doubling every five years thereafter. Among those aged 65-84, roughly one in thirteen has AD, while the number jumps to one in three for those over 85 [61]. Early-onset AD, affecting those under 65, remains rare, representing less than 10% of all cases [62]. The presence of the apolipoprotein E (APOE-*ε*4) allele, a variant of the apolipoprotein E gene, substantially increases the risk of AD in women compared to men [63, 64]. Women in their sixties who carry one or two copies of this gene variant are more likely to develop AD than their male counterparts [65].

### 4.3. Medications for AD

AD is incurable, but medications can provide relief by managing symptoms or potentially slowing disease progression. AD progressively damages and destroys nerve cells in the brain, leading to memory loss, reasoning impairment, and other cognitive declines. The objective is to decelerate this cognitive decline, enhancing the quality of life for individuals with Alzheimer’s. Here’s an overview of the two primary categories of drugs [66]:

#### 4.3.1. Symptomatic Medications

These medications are designed to alleviate symptoms such as memory loss and confusion. However, it’s important to note that these medicines are not approved or recommended for treating MCI.

1. **Cholinesterase Inhibitors:** These drugs prevent the breakdown of acetylcholine, a vital neurotransmitter involved in memory and learning.
  - **Donepezil (Aricept**^**®**^): Approved for all stages of AD.
  - **Rivastigmine (Exelon**^**®**^): Effective in managing mild to moderate AD symptoms.
  - **Galantamine (Razadyne**^**®**^): Approved for use in mild to moderate AD cases. While generally well-tolerated, these medications may lead to side effects such as nausea, diarrhea, loss of bladder control, muscle cramps, muscle twitching, and weight loss. Taking the medication at night may also result in vivid dreams [67].
2. **Glutamate Regulators:** These medications regulate glutamate, another neurotransmitter, to protect nerve cells from excessive activity that can harm them in AD patients.
  - **Memantine (Namenda**^**®**^): The only FDA-approved drug in this category for moderate to severe AD. It may slow cognitive decline and offer some nerve cell protection. Like cholinesterase inhibitors, Memantine is not a cure; side effects like dizziness, headache, confusion, hallucinations, agitation, and constipation can occur.

Our analysis reveals a strong association between AD and specific brain regions involved in memory, emotion, and spatial orientation. The effectiveness of these symptomatic medications may vary depending on which areas are affected in each patient. For example, cholinesterase inhibitors that boost acetylcholine levels, which affect cortical memory function [68], might benefit patients with substantial hippocampal involvement, given the hippocampus’s role in memory [69, 70]. Similarly, glutamate regulators like Memantine could be more beneficial for patients with damage in the entorhinal cortex [71], where excessive neural activity can worsen cognitive decline. Understanding these specific brain region predictors can help healthcare professionals tailor treatments to maximize benefits and manage side effects better.

#### 4.3.2. Disease-Modifying Drugs (Under Investigation)

These newer drugs aim to modify the underlying disease process of AD. They can potentially slow disease progression:

1. **Anti-amyloid Treatments:** These medications aim to reduce the accumulation of beta-amyloid plaques, a characteristic protein abnormality in AD.
  - **Lecanemab (Leqembi**^**®**^): This drug is an example of anti-amyloid treatment, working to remove beta-amyloid from the brain. It is approved by the FDA as an intravenous infusion for individuals in the early stages of AD with confirmed elevated brain amyloid levels. Beta-amyloid accumulates early in the entorhinal cortex and hippocampus, leading to cognitive deficits and synaptic dysfunction [72]. Therefore, Lecanemab targets these critical regions. However, Lecanemab is not a cure, and its long-term effects and safety are still being investigated. Potential side effects of taking Lecanemab may include brain swelling or bleeding, and tests such as PET scans or spinal taps are required before initiating treatment [73]. Due to the risk of brain swelling or bleeding (known as ARIA), patients receiving Lecanemab require regular MRI scans for monitoring. These scans are recommended before the fifth, seventh, and fourteenth infusions and after one year of treatment. Rarely, these adverse effects may lead to symptoms such as headache, confusion, dizziness, changes in vision, nausea, and difficulty walking.
  - **Kisunla (donanemab-azbt):** On July 2, 2024, the FDA approved Kisunla, another anti-amyloid treatment for AD. It is administered as an intravenous infusion every four weeks and is intended for patients with MCI or mild dementia. Clinical trials demonstrated that Kisunla significantly reduced clinical decline on various AD scales compared to placebo, including the Integrated Alzheimer’s Disease Rating Scale (iADRS) and the Clinical Dementia Rating Scale – Sum of Boxes (CDR-SB). However, there are risks associated with Kisunla, such as ARIA and infusion-related reactions. The FDA granted Kisunla Fast Track, Priority Review, and Breakthrough Therapy designations [74].

Aducanumab, once approved for the treatment of early-stage AD, was later withdrawn from the market due to concerns about its effectiveness [75]. Similarly, Lecanemab, which the FDA approved, has not shown substantial efficacy in halting the progression of the disease or reversing any associated damage [76].

Transcranial magnetic stimulation (TMS) and transcranial direct current stimulation (tDCS) are emerging as promising non-pharmacological interventions for AD. While previous clinical trials have primarily focused on evaluating their effects on global cognition, the authors in [77] explore the specific cognitive function of memory, which AD profoundly impacts. Using multilevel random effect models, the authors assessed the efficacy and safety of both TMS and tDCS in memory deficits among AD patients. The findings indicate that depending on the targeted brain regions, both interventions positively affected memory symptoms in AD patients. Specifically, rTMS over frontal regions and tDCS over temporal regions demonstrated improvements in memory abilities. rTMS exhibited a statistical tendency towards enhancing memory in the long term, while the prolonged effects of tDCS could not be fully assessed due to limited data. The analysis also highlighted the safety of both techniques in the AD population, with minimal reports of serious adverse events. This review suggested the potential of TMS and tDCS as viable therapeutic options for addressing memory deficits in AD patients. These findings align with our analysis, which identified the entorhinal cortex and hippocampus, both located in the temporal region, as consistently important in AD classification. The observed benefits of tDCS targeting temporal areas may reflect modulation of these same regions, reinforcing their relevance in memory-focused interventions. This anatomical convergence underscores the value of identifying predictive brain regions when evaluating non-pharmacological treatment strategies for AD.

##### Important Considerations

Our research highlights the varied neuroanatomical patterns in AD across different sex and age groups. The effectiveness of medications in slowing cognitive decline may differ among individuals based on their specific brain characteristics. For instance, regions like the hippocampus, amygdala, and entorhinal cortex are top contributors to AD and may respond uniquely to treatments. Recognizing these differences can help doctors tailor treatments to each person, maximizing benefits and reducing side effects. Therefore, seeking personalized advice from healthcare professionals is crucial to managing AD symptoms effectively. Maintaining a healthy lifestyle, including regular exercise, a balanced diet, cognitive stimulation, and good sleep hygiene, can substantially slow AD progression.

### 4.4. Challenges Faced

Several challenges were encountered during preprocessing and dataset preparation. Our analysis was constrained by the use of predefined brain atlases for parcellation. While tools like FastSurfer offer efficient and standardized segmentation, they may overlook subtle structural patterns present in raw 3D anatomical images. Deep learning approaches that operate directly on full-resolution MRI volumes could potentially capture richer spatial features and improve predictive accuracy. Additionally, the absence of complementary modalities such as functional data or gray and white matter maps limited the scope of our analysis. Expanding both the size and modality diversity of the dataset may enhance the robustness and generalizability of AD prediction models.

### 4.5. Future Work

Future work can focus on optimizing diagnostic tools for AD by integrating advanced machine learning techniques such as Randomized Search and Recursive Feature Elimination to improve model accuracy and efficiency. Deep learning methods, when combined with traditional classifiers, may enhance precision in identifying AD-related brain regions. Additionally, exploring the link between anxiety and AD could offer insights into early intervention strategies.

Other important subgroups in Alzheimer’s research that warrant further investigation include genetic variants, which can help explain how specific mutations or polymorphisms affect AD risk and progression. Exploring the impact of ethnicity and race can reveal differences in the prevalence and outcomes of disease in diverse populations. Socioeconomic factors such as education, income, and access to healthcare also influence the incidence and severity of AD. Furthermore, examining comorbidity profiles can shed light on how other medical conditions interact with AD pathology and influence disease progression.

Finally, integrating multimodal approaches such as functional near-infrared spectroscopy (fNIRS) with MRI could provide a more comprehensive understanding of brain health. However, challenges related to data integration, standardization, and computational cost must be addressed to make such approaches clinically viable.

In conclusion, we processed images from the ADNI database using FreeSurfer and applied the Random Forest algorithm with K-fold, Stratified K-fold, and Leave-One-Out cross-validation techniques to predict AD, MCI, and CN, achieving an average accuracy of 92.87% in detecting AD. Our subgroup analyses revealed that the hippocampus, amygdala, and entorhinal cortex are key predictors of Alzheimer’s across both younger (69-76) and older (77-84) age groups. Additionally, distinct brain volume reductions were observed, with the left middle temporal cortex more affected in females and the right entorhinal cortex in males. These results underscore the importance of targeted interventions and early detection of AD-related neurodegenerative changes.

## 5. Supplementary Material

### S1 - List of Brain Regions

95 brain regions of FastSurfer’s ‘segmentation-only’ pipeline were used for training the AD classifier.

#### Cortical volumes

ctx-lh-caudalanteriorcingulate, ctx-lh-caudalmiddlefrontal, ctx-lh-cuneus, ctx-lh-entorhinal, ctx-lh-fusiform, ctx-lh-inferiorparietal, ctx-lh-inferiortemporal, ctx-lh-isthmuscingulate, ctx-lh-lateraloccipital, ctx-lh-lateralorbitofrontal, ctx-lh-lingual, ctx-lh-medialorbitofrontal, ctx-lh-middletemporal, ctx-lh-parahippocampal, ctx-lh-paracentral, ctx-lh-parsopercularis, ctx-lh-parsorbitalis, ctx-lh-parstriangularis, ctx-lh-pericalcarine, ctx-lh-postcentral, ctx-lh-posteriorcingulate, ctx-lh-precentral, ctx-lh-precuneus, ctx-lh-rostralanteriorcingulate, ctx-lh-rostralmiddlefrontal, ctx-lh-superiorfrontal, ctx-lh-superiorparietal, ctx-lh-superiortemporal, ctx-lh-supramarginal, ctx-lh-transversetemporal, ctx-lh-insula, ctx-rh-caudalanteriorcingulate, ctx-rh-caudalmiddlefrontal, ctx-rh-cuneus, ctx-rh-entorhinal, ctx-rh-fusiform, ctx-rh-inferiorparietal, ctx-rh-inferiortemporal, ctx-rh-isthmuscingulate, ctx-rh-lateraloccipital, ctx-rh-lateralorbitofrontal, ctx-rh-lingual, ctx-rh-medialorbitofrontal, ctx-rh-middletemporal, ctx-rh-parahippocampal, ctx-rh-paracentral, ctx-rh-parsopercularis, ctx-rh-parsorbitalis, ctx-rh-parstriangularis, ctx-rh-pericalcarine, ctx-rh-postcentral, ctx-rh-posteriorcingulate, ctx-rh-precentral, ctx-rh-precuneus, ctx-rh-rostralanteriorcingulate, ctx-rh-rostralmiddlefrontal, ctx-rh-superiorfrontal, ctx-rh-superiorparietal, ctx-rh-superiortemporal, ctx-rh-supramarginal, ctx-rh-transversetemporal, ctx-rh-insula.

#### Subcortical gray matter volumes

Left-Cerebellum-Cortex, Left-Thalamus, Left-Caudate, Left-Putamen, Left-Pallidum, Brain-Stem, Left-Hippocampus, Left-Amygdala, Left-Accumbens-area, Left-VentralDC, Right-Cerebellum-Cortex, Right-Thalamus, Right-Caudate, Right-Putamen, Right-Pallidum, Right-Hippocampus, Right-Amygdala, Right-Accumbens-area, Right-VentralDC.

#### Ventricular system volumes

Left-Lateral-Ventricle, Left-Inf-Lat-Vent, 3rd-Ventricle, 4th-Ventricle, CSF, Left-choroid-plexus, Right-Lateral-Ventricle, Right-Inf-Lat-Vent, Right-choroid-plexus.

#### Subcortical white matter volumes

Left-Cerebral-White-Matter, Left-Cerebellum-White-Matter, Right-Cerebral-White-Matter, Right-Cerebellum-White-Matter, WM-hypointensities.

### S2 - Performance metrics and Top Contributing Features of nine sub-groups

Tables 8, 13, and 14 display the performance metrics and top contributing features obtained through K-fold, stratified K-fold, and leave-one-out cross-validation techniques.

**Table 13:**
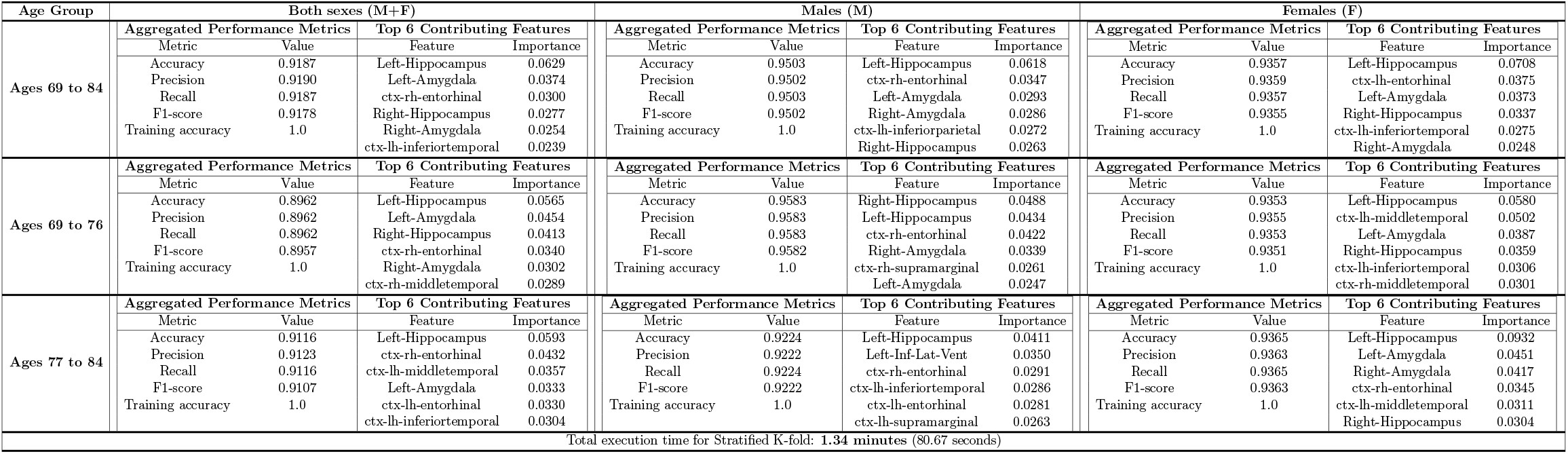
Comparison of Performance Metrics and Top Contributing Features using Stratified K-fold Cross-Validation.

**Table 14:**
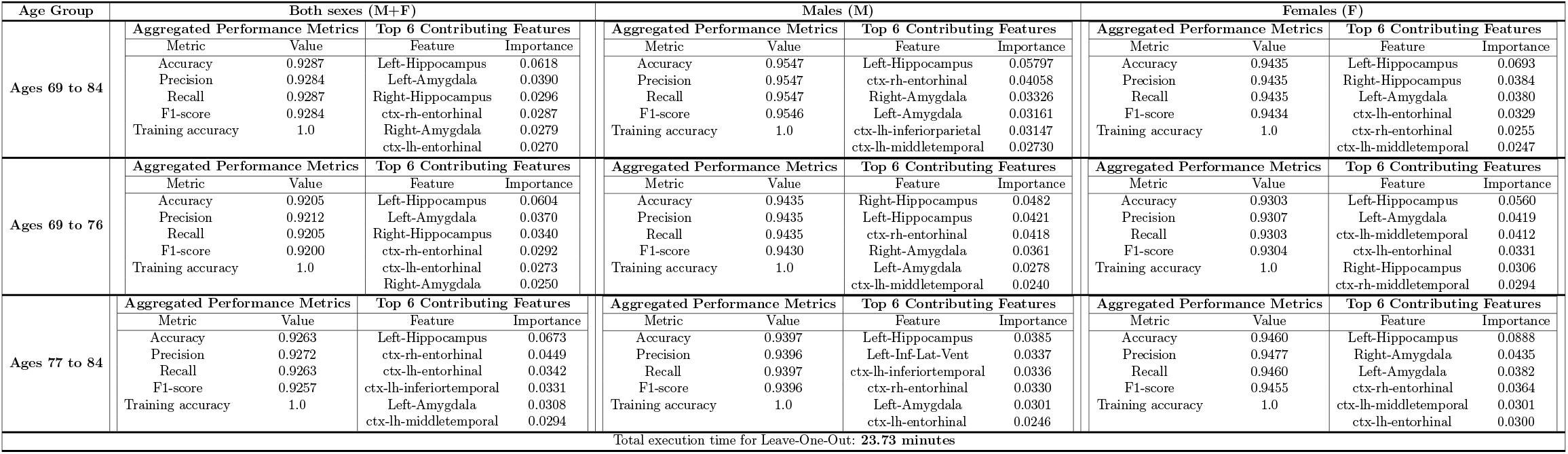
Comparison of Performance Metrics and Top Contributing Features using Leave-One-Out Cross-Validation.

## Data Availability

This study used data from the Alzheimer's Disease Neuroimaging Initiative available online upon registration at

https://adni.loni.usc.edu/

